# Histological triage of early-stage mycosis fungoides using a weakly supervised deep learning-based model: a multicentre, external validation, and clinical utility study

**DOI:** 10.64898/2026.07.27.26359009

**Authors:** Thom Doeleman, Siemen Brussee, Pieter A. Valkema, Werner Kempf, Maarten H. Vermeer, Jesper Kers, Liliane C.D. Wynaendts, Kelly G.P. Kerckhoffs, Marthe M. de Jonge, Anh H. Nguyen, Elke E.M. Peters, Marion Wobser, Hilka Rauert-Wunderlich, Andreas Rosenwald, Rudolf Stadler, Patty M. Jansen, Maxime Battistella, Gabriele Roccuzzo, Pietro Quaglino, Anne M.R. Schrader, all CLIDIPA Registry participating centres

## Abstract

**Background:** Histological diagnosis of early-stage mycosis fungoides (MF) is hindered by profound overlap with benign inflammatory dermatoses (BIDs), leading to diagnostic delays and extensive ancillary testing. We developed MIMIC (Multiple Instance-learning for Identification of Mycosis fungoides In Cutaneous biopsies), a weakly supervised deep learning model designed as a triage tool at initial HCE whole-slide image (WSI) review to distinguish classic patch- and plaque-stage MF from BIDs. We externally validated the model and evaluated its clinical utility.

**Methods:** In this retrospective multicentre study, we trained a base model using weakly supervised attention-based multiple-instance learning on 3,339 WSIs from two Dutch centres. Crucially, all MF training labels were derived from a deeply phenotyped national cohort featuring strict multidisciplinary expert panel consensus diagnoses (the clinical gold standard). Transportability was evaluated on 371 WSIs from four independent European centres. A blinded reader study on 171 WSIs compared morphology-only performance of MIMIC with 11 (dermato-)pathologists. We then retrained an updated model on all retrospective multicentre data and assessed clinical utility in a strictly held-out, consecutive Utrecht cohort (2022-2023; 486 accessions, 863 WSIs). Primary analysis focused on classic MF versus BIDs (453 accessions). Decision curve analysis, using Platt-scaled probabilities to correct for spectrum bias, evaluated net benefit at a prespecified, safety-oriented threshold of 0.04.

**Findings:** The base model showed good multicentre transportability (mean centre-specific AUROC 0.91; pooled AUROC 0.84). In the reader study, MIMIC achieved an AUROC of 0.87, exceeding the mean pathologist AUROC (0.79) and the best individual reader (0.83). In the consecutive MF versus BID cohort, the updated model achieved an AUROC of 0.87 (95% CI 0.81-0.92). At the 0.04 threshold, sensitivity was 97.8% (44/45 MF cases) and specificity 50.2%, reducing unnecessary ancillary workups by 39.9 per 100 screening cases versus a test-all strategy.

**Interpretation:** By identifying nearly half of BIDs as low risk while preserving near-complete sensitivity for classic early-stage MF in a European digital pathology workflow, this unimodal HCE approach offers a scalable digital solution to reduce defensive ancillary testing and accelerate the diagnostic journey for patients with MF. Further validation is needed in non-European centres and in populations with darker skin phototypes.

**Funding:** This work was supported by an unrestricted Fellowship grant of Stichting Hanarth Fonds in The Netherlands.

**Research in context:** *Evidence before this study:* Mycosis fungoides (MF) is notoriously difficult to diagnose in its early stages due to significant clinical and histopathological overlap with benign inflammatory dermatoses (BIDs), resulting in a median diagnostic delay of 36 months from symptom onset. Standard diagnostic workups are highly resource-intensive and frequently inconclusive, often requiring multiple consecutive biopsies, extensive immunohistochemistry panels, molecular T-cell receptor clonality studies, and iterative clinicopathological correlation in multidisciplinary consensus meetings. We systematically searched PubMed from database inception up to May 28, 2026, using the terms (“mycosis fungoides” OR “cutaneous T-cell lymphoma” OR “CTCL”) AND (“deep learning” OR “machine learning” OR “artificial intelligence” OR “AI” OR “foundation model”) with no language restrictions, identifying 123 raw citations. Only eight original research articles were found to explicitly evaluate machine learning or artificial intelligence applications for cutaneous lymphomas. This pool was supplemented with international conference proceedings and a preprint, identifying a total of thirteen relevant studies. The vast majority of this limited literature focused on non-histological modalities (e.g., clinical photography, dermoscopic imaging, 3D skin tumour burden scoring, or structured electronic health records) or required specialized, non-standard imaging hardware such as non-linear optical microscopy. Furthermore, alternative computational pipelines relied on highly resource-intensive and costly molecular data, such as bulk RNA-sequencing gene-expression classifiers. Crucially, fewer than five peer-reviewed studies focused on standard brightfield haematoxylin and eosin (HCE) whole-slide images (WSIs) to differentiate early-stage MF from BIDs. While recent baseline architectures, exploratory conference abstracts, and emerging multimodal systems integrating histopathology with clinical metadata have established the existence of a trainable discriminative signal, severe translational limitations persist. Most existing frameworks relied on small development datasets, lacked rigorous external geographic validation across independent international registries, and failed to strictly isolate patient-level clustered data during performance evaluation. Crucially, no prior study has evaluated a unimodal brightfield HCE pathology foundation model within a formalized, Platt-scaled calibration framework to correct for spectrum bias, or utilized decision curve analysis to demonstrate safe, high-sensitivity clinical triage in a consecutive screening pipeline.

*Added value of this study:* We developed, externally validated, and evaluated the clinical utility of MIMIC, a clinical decision support tool utilizing a pathology foundation model (H-Optimus-1) for risk-stratified triage at the point of initial histological review. In an external geographic validation across four independent European institutions, the base model demonstrated high transportability with a mean centre-specific AUROC of 0·91. In a blinded multi-centre reader study on 171 WSIs, MIMIC achieved an AUROC of 0·87, outperforming the mean baseline performance of 11 independent (dermato-)pathologists (AUROC 0·79 ± 0·03) and exceeding the integrated discriminative area of the top-performing human expert (AUROC 0·83). Crucially, following TRIPOD+AI guidelines for model updating, the final candidate was evaluated on a strictly held-out, consecutive clinical screening workflow cohort (*N* = 453 accessions). Following Platt scaling to robustly calibrate for spectrum bias, MIMIC achieved a diagnostic sensitivity of 97·8% at a conservative, safety-oriented operational threshold of 0·04, intercepting 44 out of 45 true malignant cases while effectively identifying 50·2% of BIDs as low-risk on initial HCE morphology alone.

*Implications of all the available evidence:* MIMIC can be seamlessly integrated into digital pathology workflows to safely “de-bulk” the diagnostic workload by reliably identifying low-risk cases that can be triaged away from automatic ancillary testing. Our automated three-tier workflow analysis demonstrates that a staggering 45·5% of the total diagnostic screening volume can be safely classified as low-risk triage zones where ancillary testing could be avoided, translating to substantial direct laboratory cost savings. Decision curve analysis demonstrated a consistent positive net benefit at the 0·04 threshold, corresponding to a net reduction of 39·9 unnecessary ancillary workups and expert multidisciplinary reviews per 100 screening cases relative to a standard “test all” clinical strategy. By acting as a highly sensitive, objective, and reproducible filter at the earliest stage of histopathological review, this unimodal HCE approach offers a scalable, zero-marginal-cost digital solution to mitigate defensive diagnostic reflexes, optimize specialized laboratory resource allocation, and significantly accelerate the diagnostic pathway for patients with early-stage MF.

## Introduction

Mycosis fungoides (MF), the most common cutaneous T-cell lymphoma (CTCL), is notoriously difficult to diagnose in its early stages due to its profound clinical and histological overlap with benign inflammatory dermatoses (BIDs). This “great imitator” status^1^ results in a median diagnostic delay of 36 months from symptom onset^2^, with the PROCLIPI registry reporting such delays in 86% of patients, exposing them to inappropriate treatments and the risk of disease progression. Furthermore, the histological subtlety of early-stage MF (eMF), characterized by sparse neoplastic T-cell infiltrates with focal epidermotropism, leads to significant diagnostic instability, with interobserver variation reported in approximately 20% of cases, even among specialists^3^.

These diagnostic challenges impose a dual burden: distress for the patient and a substantial diagnostic workload for pathology and dermatology departments. The current reliance on comprehensive clinicopathological correlation, often requiring multiple biopsies, serial immunohistochemistry (IHC), and T-cell receptor (TCR) clonality studies for every suspicious lesion, is resource-intensive and frequently yields equivocal results in early-stage disease due to the scarcity of neoplastic cells. Cutaneous TCR clonality is confirmed in only 64% of early-stage patients, and the detection of aberrant pan-T-cell marker loss carries a sensitivity as low as 10%, rendering reflexive ancillary testing in low-suspicion cases highly inefficient^2–4^. Consequently, there is an urgent need for objective diagnostic tools that can facilitate earlier recognition and reliable probabilistic triage at this critical bottleneck.

Our previous work^5^ established the existence of a trainable, discriminative deep learning (DL) signal for binary MF versus BID classification using only HCE-stained WSIs. Building upon this foundation, we developed MIMIC (Multiple Instance-learning for Identification of Mycosis fungoides In Cutaneous biopsies), a clinical decision support tool based on H-Optimus-1, a pathology-specific foundation model pretrained exclusively on data independent of all cohorts used in this study, designed for diagnostic triage at the point of initial histological review. Unlike previous frameworks constrained by individual diagnostic bias, MIMIC was developed exclusively using strict multidisciplinary expert panel consensus labels (established by the Dutch Cutaneous Lymphoma Group). This provides a highly robust clinical gold standard that effectively mitigates the significant interobserver variability historically plaguing computational models in this domain.

The present study aims to translate this foundational work into a clinically deployable tool through a phased validation and updating framework. First, a base model was developed on a deeply phenotyped retrospective dataset (Leiden and Utrecht) featuring strict expert panel consensus labels. To establish fundamental robustness, this model was subsequently evaluated across four independent European tertiary centres (Zürich, Minden, Turin, and Würzburg). None of these centres contributed to model training, thereby confirming external transportability across inevitable institutional domain shifts, including inter-laboratory variations in pre-analytical tissue processing, whole-slide scanner hardware, and local referral-driven benign case-mixes. Second, in accordance with TRIPOD+AI guidelines for model updating, the model was retrained on all available retrospective multicentre data to maximise exposure to morphological heterogeneity, yielding the final candidate.

To evaluate its intended clinical context of use, this updated model was applied to a strictly held-out test set of consecutive patch-and plaque-stage biopsies presenting with a clinical or histological suspicion of eMF (Utrecht 2022–2023). Because this institution utilises a fully digitised primary diagnostic workflow, evaluating this unselected cohort perfectly mirrors the routine, prospective triage of all incoming suspicious cases at a typical tertiary referral centre.

The clinical utility of MIMIC is evaluated through Decision Curve Analysis on this clinical utility cohort, quantifying the net benefit of a MIMIC-assisted probabilistic triage strategy, in which low-suspicion cases are triaged away from reflex IHC and TCR clonality studies, against current standard of care across a pre-specified threshold range of 1–10%. This reflects a clinical consensus where the severe harms of missing an early lymphoma (diagnostic delay) significantly outweigh the financial and temporal costs of unnecessary ancillary testing.

## Methods

### Study design and oversight

This retrospective multicentre study was designed to develop and validate the DL diagnostic prediction model MIMIC. The system was designed as a clinical decision support tool and risk-stratification aid for the early identification of MF in cases with clinical and/or histopathological overlap with BIDs. Following the framework for external validity in biomedical research and TRIPOD+AI reporting guidelines^6,7^, we conducted a phased validation and model updating strategy. First, a base model assessed the transportability of MIMIC across European centres with cutaneous lymphoma expertise to evaluate performance under laboratory-specific domain shifts. Second, in accordance with TRIPOD+AI guidelines for model updating, the algorithm was updated to maximise data exposure before evaluating its clinical utility on a strictly held-out, consecutive clinical workflow cohort (Utrecht 2022–2023). The complete TRIPOD+AI checklist is provided in the supplements.

### Cohorts

#### Base Model Development and External Geographic Validation

The development of the base model utilised a pathologist-curated dataset from two national centres: Leiden University Medical Centre and University Medical Centre Utrecht. This cohort included 2,642 WSIs from 1,887 patients diagnosed between March 2010 and September 2024. External transportability of this frozen base model was evaluated on 371 WSIs sourced from four international centres within the Cutaneous Lymphoma International Digital Pathology (CLIDIPA) Registry: Zürich (Switzerland), Turin (Italy), Würzburg (Germany), and Minden (Germany).

#### Model Updating and Held-out Clinical Workffow Validation

To maximise the training signal and exposure to morphological heterogeneity prior to clinical utility evaluation, the algorithm was subsequently updated. This updated deployment model was retrained on the cumulative dataset from all participating centres (Leiden, Utrecht, and the four international sites), encompassing available retrospective data up to September 2024.

Crucially, to ensure a robust and unbiased evaluation of real-world clinical utility, a distinct evaluation cohort (N=486 cases) was strictly held out from all model development and updating phases. This held-out validation cohort comprised every consecutive patient with a clinical and/or histological suspicion of eMF at Utrecht between 1 January 2022 and 31 December 2023. Cases were identified via the Laboratory Information System (Delphic AP, Sysmex) using a standardised search strategy (see Appendix) designed to capture the clinical diagnostic bottleneck. Inclusion required either a clinical request explicitly mentioning “rule-out MF” or a pathologist-initiated request for T-cell IHC profiling due to morphological suspicion of MF on initial HCE review. Because this held-out cohort represents a chronologically consecutive, unmanipulated clinical workflow of suspicious patch- and plaque-stage lesions, it rigorously reflects the true clinical case-mix and natural disease prevalence required to accurately evaluate the model’s downstream clinical utility.

### Reference standard and ground truth definition

For the Dutch development datasets (Leiden and Utrecht), MF diagnoses were established through the registry of the Dutch Cutaneous Lymphoma Group (DCLG). The DCLG is a national multidisciplinary collaboration of dermatologists and pathologists from all Dutch university medical centres, meeting 4 times annually to establish consensus on new patients with suspected cutaneous lymphomas. Final diagnoses were synthesized in accordance with the WHO-EORTC classification following a comprehensive review of clinical photography, longitudinal clinical follow-up, and representative skin biopsies, integrating advanced ancillary testing where applicable. For the external CLIDIPA centres (Zürich, Würzburg, Minden, and Turin), primary histopathological evaluation was performed by board-certified (dermato-)pathologists and subspecialist CTCL experts. Final diagnoses integrated clinical presentation, histopathology, and routine ancillary studies, including comprehensive IHC phenotyping and, when indicated, TCR clonality studies.

For the control group comprising BIDs, ground truth was established based on standard-of-care clinicopathological evaluation at the time of the initial biopsy. Reflecting routine, real-world dermatopathology workflows, the extent of ancillary testing was inherently tailored to the degree of morphologic suspicion. In cases where the initial HCE evaluation revealed a clear benign inflammatory pattern lacking an atypical lymphocytic substrate, diagnoses were confidently rendered without reflexive ancillary studies to avoid unnecessary testing. Conversely, cases presenting with persistent histological ambiguity underwent a comprehensive diagnostic workup, incorporating extended IHC panels, TCR clonality studies, and/or multidisciplinary expert consultation, to definitively rule out a malignant process. All reference standard diagnoses were established as part of routine clinical care prior to this study, entirely blinded to the predictions generated by the AI model. To preserve the integrity of the benign ground-truth labels, regardless of the initial diagnostic workup depth, and minimise the risk of including patients with undiagnosed, early-evolving MF, a robust longitudinal follow-up was performed. Patient trajectories within the clinical utility cohort were verified up to February 2026 by reviewing local electronic health records and the Laboratory Information System for subsequent biopsies. Furthermore, to capture potential out-of-network progression, cases were cross-referenced against the nationwide DCLG registry. Because the DCLG operates under strict privacy regulations, linkage was performed using available pseudo-anonymised demographic identifiers (date of birth, sex, and initials). While this approach precludes absolute deterministic linkage, it substantially mitigates the likelihood of missing subsequent MF diagnoses.

### Development phase (model training)

For the Dutch training cohorts, MF patients were retrospectively included if they were diagnosed via the DCLG consensus between March 11, 2010, and September 12, 2024. Representative skin biopsies from all patients with early stages of classic MF (stages IA–IIA) and only patch-or plaque-stage disease from patients with advanced stages of classic MF (stages IIB–III) were included. We excluded biopsies with tumour-stage disease and transformed large-cell variants. Furthermore, the specific variants of MF, namely folliculotropic MF (FMF), pagetoid reticulosis, and granulomatous slack skin, were excluded from the training data, as these entities present with distinct histological and clinical mimickers that differ from the diagnostic challenge posed by classic patch- and plaque-stage MF.

For the control (non-MF) cohort in the development phase, we included representative skin biopsies encompassing a broad spectrum of BIDs. This cohort comprised a mix of cases sent for expert consultation due to clinical and/or histological suspicion of MF, as well as a diverse selection of established MF mimickers. Based on prevailing dermatopathology consensus^189^, these mimickers were explicitly selected to encompass the primary histopathological reaction patterns known to exhibit profound overlap with eMF: spongiotic (e.g., eczematous dermatitis), psoriasiform (e.g., psoriasis), and interface/lichenoid dermatitis (e.g., lichen planus, cutaneous lupus erythematosus). Furthermore, to ensure algorithmic robustness against the extensive morphological heterogeneity encountered in real-world diagnostic workflows, the control dataset was deliberately expanded to include a wider variety of inflammatory patterns, such as granulomatous, vesiculobullous, and pseudolymphomatous dermatoses. A comprehensive breakdown of the specific BID diagnoses included in the development cohort is provided in Supplementary Table 1. Consistent with standard diagnostic practice, biopsies exhibiting unequivocally benign histopathological features by the reviewing pathologist upon initial HCE review were diagnosed without the requirement for extensive ancillary testing, even if eMF was part of the clinical differential diagnosis. Conversely, in cases presenting with histopathological ambiguity, advanced ancillary testing (including IHC and TCR clonality analysis) was utilized to support exclusion of eMF.

### Clinical utility evaluation

In contrast to the curated training data, the 2022–2023 clinical utility cohort followed a search-driven inclusion policy to reflect the specific clinical challenge where an objective “rule-out MF” decision is required. This cohort reflects an unmanipulated clinical workflow by including all biopsies presenting as macules, papules, patches, or plaques where a clinical or histological suspicion of early CTCL was raised. Following the established diagnostic scope, all cases presenting as nodules or tumours were excluded. Due to the search criteria, the resulting set included classic MF as well as related variants encountered in routine practice, such as adnexotropic (folliculotropic) MF and lymphomatoid papulosis (LyP) presenting with patch- or plaque-like clinical features. Detailed inclusion and exclusion criteria based on clinical presentation and diagnostic workup are provided in the Supplementary Appendix.

### Expert Reader Study (Benchmark)

To benchmark the AI system against current clinical standards and gauge the intrinsic diagnostic difficulty of the validation cases, a multi-centre reader study was conducted utilizing a stratified random subset of 171 WSIs. To capture variations in geographic case mix and slide preparation while ensuring a balanced representation of diagnoses, this benchmark cohort was constructed to include 121 external WSIs (Zürich, n=47; Minden, n=49; Würzburg, n=25) and 50 internal WSIs (Utrecht).

Crucially, to prevent data leakage and ensure an unbiased performance evaluation in accordance with PROBAST+AI methodological criteria ^10^, the 50 Utrecht-derived WSIs were strictly sequestered and completely excluded from all model training and hyperparameter tuning phases. An independent panel of eleven (dermato-)pathologists blindly evaluated this cohort to establish the model’s comparative diagnostic accuracy relative to human specialists. The panel comprised two distinct expertise levels: six subspecialist CTCL experts (three dermatologists/dermatopathologists and three pathologists hailing from tertiary cutaneous lymphoma referral centres) and five general pathologists with an affinity for dermatopathology from general hospitals in the Netherlands.

All scoring was performed on digitized HCE-stained slides via the Slide Score online platform (https://www.slidescore.com/) using private access links. Pathologists were blinded to the ground truth diagnoses, the AI model predictions, and the assessments of other participants; they were also prohibited from scoring cases originating from their own centres. To ensure a fair, head-to-head comparison with the AI model, which operates solely on morphological pixel data, the human readers were intentionally blinded to all clinical metadata. While clinicopathological correlation is standard practice and is known to significantly improve human diagnostic accuracy and confidence in MF, this strict blinding protocol was necessary to isolate the baseline diagnostic value of the HCE morphology and prevent differential information bias.

For each WSI, (dermato-)pathologists provided a diagnostic score on a structured five-point ordinal scale: mycosis fungoides (probable), mycosis fungoides (uncertain), completely uncertain diagnosis, benign inflammatory dermatosis (uncertain), and benign inflammatory dermatosis (probable). This ordinal assessment was translated into a numerical accuracy-confidence score based on agreement with the multidisciplinary ground truth: +2 (correct and probable), +1 (correct but uncertain), 0 (completely uncertain), -1 (incorrect but uncertain), and -2 (incorrect and probable).

Based on the mean consensus score calculated across all eleven (dermato-)pathologists, the 171 cases were dichotomized by the median into two analytical cohorts: an “easy” cohort (the top 85 cases demonstrating the highest human consensus and diagnostic accuracy) and a “difficult” cohort (the remaining 86 cases with the lowest average scores).

Additionally, the panel evaluated the predominant histopathological reaction pattern for each case, selecting from spongiotic, lichenoid/interface, psoriasiform, vasculopathic, vesiculobullous, granulomatous, or non-specific. A single predominant reaction pattern for each WSI was established via majority voting among the eleven readers. Cases resulting in a tied vote, reflecting high morphological ambiguity, were classified as ’ambiguous’.

### Experimental design / development Image acquisition and digitization

For the development and internal updating of the model, HCE-stained slides from representative skin biopsies were collected from the (digitized) archives of the Department of Pathology at the Leiden University Medical Centre (LUMC) and University Medical Centre Utrecht (UMCU). This included slides from in-house biopsies as well as locally stained slides retrieved from consultation cases. To capture real-world scanner variability, the Leiden cohort was digitized at 40X magnification using three distinct scanner models: Philips Ultra-Fast Scanner (18·8%; 0·25 mpp), 3DHISTECH Pannoramic 250 Flash II (2·7%; 0·1945 mpp), and 3DHISTECH Pannoramic P480 (78·5%; 0·25 mpp). The Utrecht cohort was digitized using an Aperio ScanScope XT at 20X magnification (20·5%) and a Hamamatsu NanoZoomer S360 at 40X magnification (79·5%).

Slides for the independent external validation cohorts were retrieved from the Cutaneous Lymphoma International Digital Pathology (CLIDIPA) Registry (https://clidipa.org/), an international platform facilitating AI-based research in cutaneous lymphomas via a large, multicentre database of digital pathology images. Anonymized slides from the participating centres (Zürich, Turin, Würzburg, and Minden) were uploaded to the Slide Score digital platform or transferred directly to the LUMC coordinating centre. Depending on the originating institution, these slides introduced further hardware-induced domain shifts, having been variably scanned by Hamamatsu (40X magnification; ∼0·23 mpp), Aperio ScanScope XT (20X magnification; 0·50 mpp), and 3DHISTECH Pannoramic SCAN 150 (20X magnification; 0·2738 mpp) scanner models. All model training, cross-validation, and feature extraction pipelines were executed on dedicated computational hardware utilizing an NVIDIA RTX 6000 GPU with 48 GB of GDDR6-RAM.

### Image preprocessing and input data eligibility

In accordance with TRIPOD+AI guidelines for input data quality, strict preprocessing and eligibility criteria were applied. Out-of-focus WSIs were rescanned where possible. WSIs exhibiting severe fading of the HCE stain or completely lacking tissue were excluded. Crucially, WSIs missing the epidermal layer were excluded from the study, as the presence of the epidermis is biologically requisite for the evaluation of epidermotropism, a hallmark of eMF. Prior to analysis, WSIs in DICOM format were converted into pyramidal TIFF format using the open-source DICOMtoSVS tool (available from: https://github.com/bertrandchauveau/DICOMtoSVS). Differences in native scanner resolutions were addressed during tile extraction, where all image patches were rescaled to a fixed spatial resolution of 0·50 mpp.

To maximize the purity of the morphological signal during model development, artifact-free and representative tissue sections in the training cohorts were manually delineated to minimize redundant information from sequential sections. The annotated regions were then exported to tiled TIFF format images using Slidescape (available from https://github.com/amspath/slidescape). Conversely, to rigorously evaluate the algorithm’s real-world clinical utility, all available tissue sections were retained without manual curation for the external and held-out clinical workflow validation cohorts. This ensured the model was evaluated on its ability to autonomously navigate uncurated tissue containing routine histological artifacts.

### WSI Processing and Feature Extraction

WSIs were digitized, segmented using a Gaussian-blur-preprocessed Otsu thresholding algorithm (*σ*=2·0, *τ*=0·02) to isolate tissue from background, and tessellated into non-overlapping tiles. Based on an extensive benchmarking suite evaluating multiple pathology foundation models (see Supplementary Appendix), we selected the H-Optimus-1 foundation model^11^ as a frozen feature extractor. Tissue segmentation, tiling, and feature extraction were all performed using the open-source PathBench-MIL software package.^12^ Each WSI was processed at a spatial resolution of 0·5 μm/pixel (approximately 20× magnification) with a tile size of 256 × 256 pixels.

### Model Architecture and Training

MIMIC utilises an Attention-based Multiple Instance Learning (ABMIL)^13^ framework with gated attention pooling. To ensure standardized evaluation and reproducibility, we employed the default ABMIL architectural settings as implemented in the open-source MIL-Lab framework.^14^ During the base model development phase, a 10-fold cross-validation scheme was implemented to obtain a more reliable estimate of model generalisation and reduce potential overfitting to a specific train–test split. Models were trained for 50 epochs using a batch size of 8, utilising the AdamW optimizer with an initial learning rate of 1 × 10^−4^ and a weight decay of 1 × 10^−5^. To dynamically adjust the optimization pathway, a ReduceLROnPlateau training scheduler was employed based on validation loss monitoring, with a decay factor of 0·1, a patience of 5 epochs, and a minimum learning rate floor of 1 × 10^−7^. To achieve stable gradients, gradient accumulation was applied across every 4 training passes, yielding an effective batch size of 32 for optimization.

To mitigate structural class imbalance in the training data, the binary cross-entropy loss function incorporated inverse-occurrence-weighted class weights. Monte Carlo Dropout (100 stochastic forward passes) was used to estimate predictive uncertainty per patient, quantified via predictive variance and entropy. Following external validation, the model underwent a final updating phase using all available retrospective data, employing the optimal hyperparameters identified during cross-validation, prior to clinical utility evaluation.

### Model evaluation and statistical analysis

In accordance with the TRIPOD+AI guidelines for reporting clinical prediction models and recent consensus recommendations for machine learning in oncology^6,7^, model evaluation focused on three core dimensions of performance: discrimination, calibration, and clinical utility^15^. To mirror clinical decision-making, WSI-level predictions were aggregated to the accession level (case-level) using a max-pooling operator, defined as *p^_case_* = *max*(*p^_wsi_*_,1_, …, *p^_wsi_*_,*n*_). This approach ensures that any high-risk histological slide identified by the model triggers clinical attention for the entire case. For discrimination, we report the Area Under the Receiver Operating Characteristic Curve (AUROC). No formal a priori sample-size calculation was performed; the study size was determined pragmatically by the availability of eligible retrospective cases. For model development, 3,339 WSIs (912 MF, 2,427 BIDs) provide ample events for deep learning model training. For clinical utility, 453 consecutive accessions (45 MF cases) were considered sufficient to estimate AUROC and decision-curve net benefit with acceptable precision.

### Calibration and recalibration architecture

#### Base model calibration (Geographic Validation)

For the evaluation of the base model across the four independent external validation cohorts, post-hoc outputs were calibrated at the logit level using scalar temperature scaling. The temperature scaling parameters were optimized on the validation logits using a Limited-memory Broyden–Fletcher–Goldfarb–Shanno (L-BFGS) algorithm with a learning rate of 0·01 and a maximum of 50 iterations. These calibrated validation outputs were subsequently aggregated across the 10 folds using a temperature-weighted mean to provide the final case-level model logits.

#### Simulation of Local Recalibration and Early Deployment (Clinical Utility Cohort)

Conversely, to correct for severe spectrum bias observed in the highly suspect clinical utility cohort, we performed local model updating via Platt scaling (logistic regression fitted directly to the raw model logits, bypassing the prior temperature scaling step). To rigorously prevent patient-level data leakage during this recalibration, we employed a 5-fold stratified cross-validation scheme grouped by patient identifier. To simulate an early clinical deployment scenario, where an adopting hospital has only limited local data available for calibration, we inverted the traditional cross-validation ratio (20% calibration → 80% application). In each fold, approximately 20% of the cohort was used to fit the Platt scaling parameters, which were subsequently applied to generate probabilities for the remaining 80%. Consequently, each patient case received out-of-fold calibrated probabilities from four independent scaler models. To yield a single, stable risk estimate per case, these four out-of-fold predictions were averaged.

Calibration performance for the deployment model was assessed using smoothed calibration plots (loess) to evaluate the agreement between predicted probabilities and observed outcomes, with 95% confidence intervals estimated via 1,000 bootstrap resamples of the final averaged predictions. Furthermore, the distribution of estimated probabilities for each outcome category was visualized using violin plots. Because the primary clinical role of MIMIC is to support triage decisions, the model is used in deployment as a risk-ranking tool with prespecified thresholds, rather than as a source of fully reliable absolute risk estimates.

Clinical utility was evaluated using Decision Curve Analysis (DCA) on the strictly held-out clinical workflow cohort from Utrecht (N=486 accession numbers, comprising 863 WSIs), simulating a clinical context of use scenario at the point of initial histological review. We calculated the Net Benefit (NB) across a clinically relevant range of threshold probabilities (1-10%). This range reflects the decision-analytical trade-off between the benefit of identifying eMF and the harm/cost associated with unnecessary ancillary testing or expert referral for benign cases. The lower bound of the threshold (0·01) reflects a clinical preference that prioritises high sensitivity (minimising missed cancer diagnoses), while higher thresholds model a more conservative approach to healthcare resource allocation. By comparing the NB of the MIMIC-assisted triage strategy against the default “test all” (current standard of care) and “test none” strategies, we quantified the potential reduction in diagnostic burden. While DCA served as the primary measure of clinical utility, descriptive fixed-threshold classification metrics (sensitivity, specificity and negative predictive value) at a prespecified safety-oriented decision threshold of 0·04 are also reported to provide comprehensive clinical context.

For the human–AI reader study, agreement between MIMIC and individual pathologists was quantified using linearly weighted Cohen’s κ on a five-point ordinal diagnostic scale. Fleiss’ kappa was not calculated due to variable numbers of ratings per case, as pathologists did not evaluate slides originating from their own centre.

For demographic and categorical variables (e.g., sex, biopsy location), P-values were obtained via the Pearson chi-square test. For continuous variables with non-normal distributions (e.g., patient age, slide age), the Mann-Whitney U test was employed. All p-values below 0·05 were considered statistically significant. 95% confidence intervals were estimated using bootstrap resampling (1000 iterations). Analyses were performed using Python (version 3.12) with the Slideflow (v2.3.0) and scikit-learn libraries. Descriptive statistics and agreement measures were supplemented using IBM Statistical Package for the Social Sciences (version 28) and Microsoft Excel (version 2403).

### Code Availability

The complete pipeline for model training, feature extraction, evaluation, and the cluster-robust bootstrap analysis is fully documented and publicly available in the dedicated GitHub repository: https://github.com/LUMCPathAI/MIMIC.

### Ethics Statement

The study was performed in accordance with the Declaration of Helsinki. Written informed consent was waived owing to the retrospective nature of the study, the rarity of CLs and the extended inclusion period of over 15 years, during which many patients were no longer under care or had passed away. Patients who actively objected to the use of their medical data or residual materials were excluded. Ethical approval was granted by the non-Medical Research Involving Human Subjects Act committee of division 4 of the Leiden University Medical Centre (nWMO-D4-2024-015), and review by the Medical Ethics Committee was not required. No separate pre–registered study protocol was prepared, and the study was not prospectively registered in a clinical trial or observational study registry. Patients and members of the public were not involved in the design, conduct, reporting, or dissemination of this study.

## Results

### Baseline dataset characteristics

A total of 4,623 whole-slide images (WSIs) from six European centres were included. The base-model development cohort comprised 2,642 WSIs (791 MF; 1,851 BIDs) from Leiden University Medical Centre and 697 WSIs (121 MF; 576 BIDs) from University Medical Centre Utrecht for a total of 3,339 WSIs. An additional 50 selected WSIs (25 MF; 25 BIDs) from Utrecht were excluded from training for inclusion in the reader study.

In the MF training subset, the median age at biopsy was 61·0 years (SD ±17·1), and 63·8% of patients were male (of known sex). The BID training subset showed a median age of 56·0 years (SD ±18·3), with 47·4% of patients being male (of known sex). Notably, Fitzpatrick skin type was undocumented for the majority of the development cohort (>97%). Technical diversity was high in the training set, utilizing five different scanner types.

The external geographic validation cohort included 371 WSIs (165 MF; 206 BID) from Zürich, Turin, Würzburg, and Minden. In contrast to the training data, clinical records in the geographic validation cohort featured substantially higher documentation completeness regarding Fitzpatrick skin type, with explicit entries for Type II (40·7% to 41·7%) and Type III (4·9% to 16·5%). While 49·7% of the geographic validation cases lacked an explicit TNM stage in the digital record, clinical documentation confirmed that all such cases presented with patch or plaque-stage disease. Detailed clinicopathological characteristics and scanner distributions for all cohorts are summarized in Table 1.

**Table 1:** Baseline cohort characteristics. *Only biopsies included from patch or plaque lesion in patients with advanced-stage MF (stage IIB to IVA2). For patients where the exact date of birth was unknown and only the year was reported, the birth date was imputed to be July 1st of the reported year to minimize potential age estimation bias. For slides where the exact date of the slide creation was unknown and only the year was reported, the slide creation date was imputed to be July 1st of the reported year to minimize potential age estimation bias. Median age of HCE slide, in years (range) calculated compared to 20-11-2025. Abbreviations: BID, benign inflammatory dermatosis; MF, mycosis fungoides; SD, standard deviation; WSI, whole-slide image. †The total consecutive screening workflow encompassed 486 accessions (863 WSIs), including 33 accessions (56 WSIs) presenting with non-MF CTCL variants that were excluded from the primary binary analysis but utilized for secondary exploratory utility evaluation.

|  | Development cohort<br>Base model<br>n=3339 WSIs |  | External geographic validation<br>Base model<br>n=371 WSIs |  | Clinical utility cohort<br>Updated model<br>n=807 WSIs<sup>†</sup> |  |
| --- | --- | --- | --- | --- | --- | --- |
|  | MF | BID | MF | BID | MF | BID |
| Number of patients (n) | 425 | 1993 | 139 | 182 | 35 | 364 |
| Number of WSIs (n) | 912 | 2427 | 165 | 206 | 74 | 733 |
| Median number of WSIs per patient (range) | 2 (1-10) | 1 (1-11) | 1 (1-4) | 1 (1-4) | 2 (1-5) | 2 (1-9) |
| Sex, male (n, % of known) | 270 (63.8%) | 771 (47.4%) | 84 (62.2%) | 107 (62.9%) | 21 (60.0%) | 214 (58.8%) |
| Sex, unknown (n, %) | 2 (0.5%) | 366 (18.4%) | 4 (2.9%) | 12 (6.6%) | 0 (0.0%) | 0 (0.0%) |
| Median age at biopsy, in years (±SD) | 61.0 ($\pm 17.1$) | 56.0 ($\pm 18.3$) | 64.5 ($\pm 14.9$) | 59.0 ($\pm 18.7$) | 68.0 ($\pm 18.9$) | 65.0 ($\pm 17.3$) |
| Fitzpatrick: Type I (n, %) | 0 (0.0%) | 0 (0.0%) | 4 (2.9%) | 2 (1.1%) | 0 (0.0%) | 0 (0.0%) |
| Fitzpatrick: Type II (n, %) | 5 (1.2%) | 0 (0.0%) | 58 (41.7%) | 74 (40.7%) | 1 (2.9%) | 0 (0.0%) |
| Fitzpatrick: Type III (n, %) | 1 (0.2%) | 0 (0.0%) | 23 (16.5%) | 9 (4.9%) | 0 (0.0%) | 0 (0.0%) |
| Fitzpatrick: Type IV (n, %) | 1 (0.2%) | 0 (0.0%) | 0 (0.0%) | 7 (3.8%) | 1 (2.9%) | 0 (0.0%) |
| Fitzpatrick: Type V (n, %) | 2 (0.5%) | 0 (0.0%) | 0 (0.0%) | 1 (0.5%) | 1 (2.9%) | 0 (0.0%) |
| Fitzpatrick: Type VI (n, %) | 0 (0.0%) | 0 (0.0%) | 0 (0.0%) | 0 (0.0%) | 0 (0.0%) | 0 (0.0%) |
| Fitzpatrick: Unknown (n, %) | 416 (97.9%) | 1993 (100.0%) | 54 (38.8%) | 89 (48.9%) | 32 (91.4%) | 364 (100.0%) |
| Scanner: Pannoramic 1000 (n, %) | 499 (54.7%) | 1574 (64.9%) |  |  |  |  |
| Scanner: Pannoramic 250 Flash II (n, %) | 18 (2.0%) | 53 (2.2%) |  |  |  |  |
| Scanner: Pannoramic SCAN 150 (n, %) |  |  | 93 (56.4%) | 139 (67.5%) |  |  |
| Scanner: Aperio ScanScope XT (n, %) | 16 (1.8%) | 133 (5.5%) |  |  |  |  |
| Scanner: Hamamatsu (n, %) | 105 (11.5%) | 443 (18.3%) | 72 (43.6%) | 67 (32.5%) | 74 (100.0%) | 733 (100.0%) |
| Scanner: Philips (n, %) | 274 (30.0%) | 224 (9.2%) |  |  |  |  |
| Loc: Head and neck (n, %) | 9 (1.0%) | 55 (2.3%) | 3 (1.8%) | 15 (7.3%) | 5 (6.8%) | 115 (15.7%) |
| Loc: Chest (n, %) | 3 (0.3%) | 5 (0.2%) | 12 (7.3%) | 5 (2.4%) | 0 (0.0%) | 45 (6.1%) |
| Loc: Trunk excl. chest (n, %) | 326 (35.7%) | 188 (7.7%) | 45 (27.3%) | 45 (21.8%) | 33 (44.6%) | 240 (32.7%) |
| Loc: Upper extremity (n, %) | 112 (12.3%) | 138 (5.7%) | 20 (12.1%) | 28 (13.6%) | 20 (27.0%) | 135 (18.4%) |
| Loc: Lower extremity (n, %) | 159 (17.4%) | 143 (5.9%) | 34 (20.6%) | 49 (23.8%) | 13 (17.6%) | 136 (18.6%) |
| Loc: Buttock/groin/genital area (n, %) | 36 (3.9%) | 9 (0.4%) | 29 (17.6%) | 7 (3.4%) | 2 (2.7%) | 49 (6.7%) |
| Loc: Unknown (n, %) | 267 (29.3%) | 1889 (77.8%) | 22 (13.3%) | 57 (27.7%) | 1 (1.4%) | 13 (1.8%) |
| Median age of H&E slide, in years (range) | 6.0 (1.0-25.1) | 10.6 (1.9-20.5) | 7.8 (0.8-17.0) | 9.7 (0.4-15.4) | 3.0 (1.9-3.9) | 2.8 (1.9-3.9) |
| Stage: IA (n, %) | 453 (49.7%) | - | 41 (24.8%) | - | 17 (23.0%) | - |
| Stage: IB (n, %) | 363 (39.8%) | - | 28 (17.0%) | - | 19 (25.7%) | - |
| Stage: IA/B (n, %) | 10 (1.1%) | - | 0 (0.0%) | - | 0 (0.0%) | - |
| Stage: IB/II (n, %) | 4 (0.4%) | - | 0 (0.0%) | - | 0 (0.0%) | - |
| Stage: IIA (n, %) | 1 (0.1%) | - | 1 (0.6%) | - | 0 (0.0%) | - |
| Stage: IIB* (n, %) | 16 (1.8%) | - | 8 (4.8%) | - | 4 (5.4%) | - |
| Stage: III* (n, %) | 0 (0.0%) | - | 3 (1.8%) | - | 5 (6.8%) | - |
| Stage: IVA1* (n, %) | 0 (0.0%) | - | 1 (0.6%) | - | 0 (0.0%) | - |
| Stage: IVA2* (n, %) | 0 (0.0%) | - | 1 (0.6%) | - | 0 (0.0%) | - |
| Stage: Unknown* (n, %) | 65 (7.1%) | - | 82 (49.7%) | - | 29 (39.2%) | - |

#### Clinical utility evaluation cohort

The consecutive clinical utility cohort represented a real-world clinical workflow at Utrecht (2022–2023), consisting of 419 patients, 486 accessions (cases), and 863 WSIs. This cohort included 74 MF WSIs, 733 BID WSIs, and 56 WSIs from other cutaneous T-cell lymphoma (CTCL) entities. The clinical utility cohort was by definition characterized by significantly more recent tissue samples compared to the historical training data, with a median HCE slide age of 2·8 to 3·0 years (range 1·9–3·9). Furthermore, this cohort represented a complete technical shift, utilizing exclusively Hamamatsu scanners (100%). Within this cohort, the accession-level operational prevalence of CTCL was 16·0% (n=78 cases), including classic MF (9·3%; n=45 cases) and other CTCL entities (6·8%; n=33 cases). Demographics remained consistent with the training data, with median ages of 65·0 to 68·0 years and a male predominance (58·8% to 60·0%).

### Base model: geographic external validation (WSI level-evaluation)

The base model, trained on retrospective data from Leiden and Utrecht, was evaluated at the WSI level across four independent external cohorts. Discriminative performance was consistent across centres, with a mean centre-specific AUROC of 0·91. Performance was highest in the Würzburg cohort (AUROC 0·96, 95% CI 0·87–1·00; n=29 WSIs), followed by Minden (0·92, 0·87– 0·97; n=87), Turin (0·90, 0·79–0·97; n=48), and Zürich (0·85, 0·80–0·90; n=196) (Figure 2A). In pooled external analysis, the AUROC was 0·84 (95% CI 0·80–0·88). This lower pooled estimate relative to the high centre-specific performance reflects expected heterogeneity in case-mix composition and baseline diagnostic difficulty across the institutional cohorts, despite preserved within-centre discrimination.

**Figure 1.**
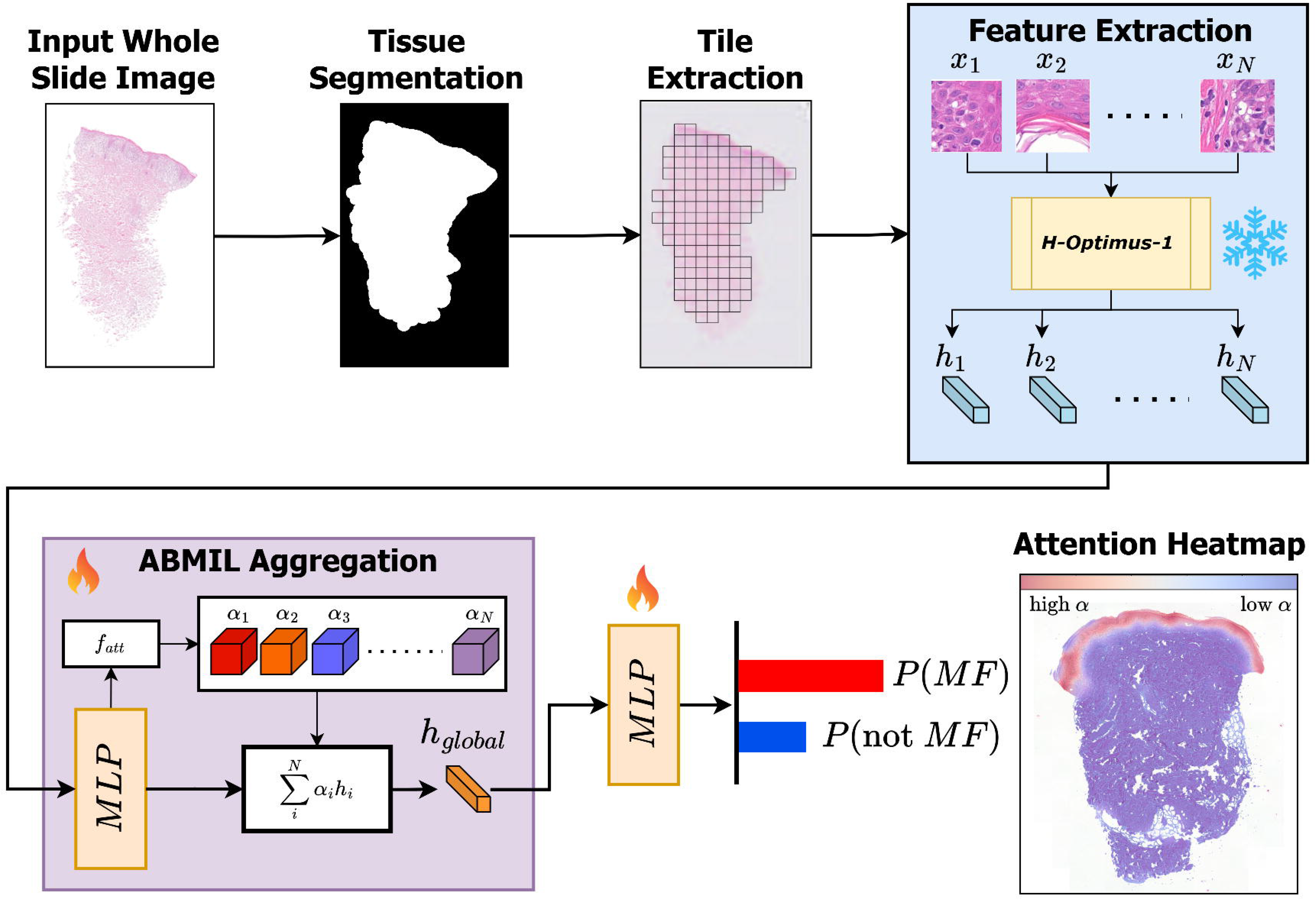
Overview of the MIMIC multiple-instance learning pipeline for automated detection of mycosis fungoides. (A) Schematic overview of the MIMIC (Multiple Instance learning for Identification of Mycosis fungoides In Cutaneous biopsies) computational pathology pipeline for whole-slide image (WSI)-based risk stratification of mycosis fungoides (MF). Haematoxylin and eosin (HCE)-stained WSIs first undergo tissue segmentation to exclude non-informative background regions. The segmented tissue area is subsequently divided into smaller image tiles for downstream analysis. Each tile is processed independently using the frozen H-Optimus-1 pathology foundation model, which extracts high-dimensional feature embeddings (ℎ_1_ … ℎ*_N_*) representing local histomorphological patterns. Tile-level embeddings are aggregated using attention-based multiple-instance learning (ABMIL), in which an attention network (*f_att_*) assigns attention weights (*α*) to diagnostically relevant regions. Weighted aggregation produces a slide-level latent representation (ℎ*_global_*), which is subsequently classified by a multilayer perceptron (MLP) to generate final probabilities for MF versus non-MF. The attention mechanism additionally enables generation of interpretable attention heatmaps highlighting tissue regions that contributed most strongly to the model prediction. Red regions indicate high attention weights corresponding to diagnostically informative areas, whereas blue regions indicate low-attention regions with limited contribution to classification. The overall framework allows slide-level prediction without requiring manual region-level annotation. (B) Study design, dataset composition, and model development workflow. Overview of dataset composition and the sequential development and evaluation strategy for the MIMIC framework. The base model was initially trained on retrospective WSIs from Leiden and Utrecht (n=3339 WSIs), comprising MF and BIDs. External validation was subsequently performed on four independent European cohorts from Minden, Turin, Würzburg, and Zürich (total n=371 WSIs). Following external validation, the updated model was retrained using all available retrospective multicentre data (n=3715 WSIs) prior to clinical utility evaluation. Following Platt scaling to robustly calibrate for spectrum bias, clinical utility was assessed in a strictly held-out consecutive Utrecht clinical utility cohort (2022–2023) consisting of cases with clinical and/or histological suspicion of early-stage MF (807 WSIs aggregated into 453 pathology accessions). * For updated model retraining, 50 additional Utrecht WSIs originating from the blinded reader study were incorporated because they were not included in either the original base-model training cohort or the external validation datasets. Conversely, 45 WSIs that were originally part of the base-model training cohort were excluded from updated model retraining because these cases were reassigned to the held-out clinical utility cohort. This reassignment was performed to prevent data leakage between model development and clinical utility evaluation.

**Figure 2.**
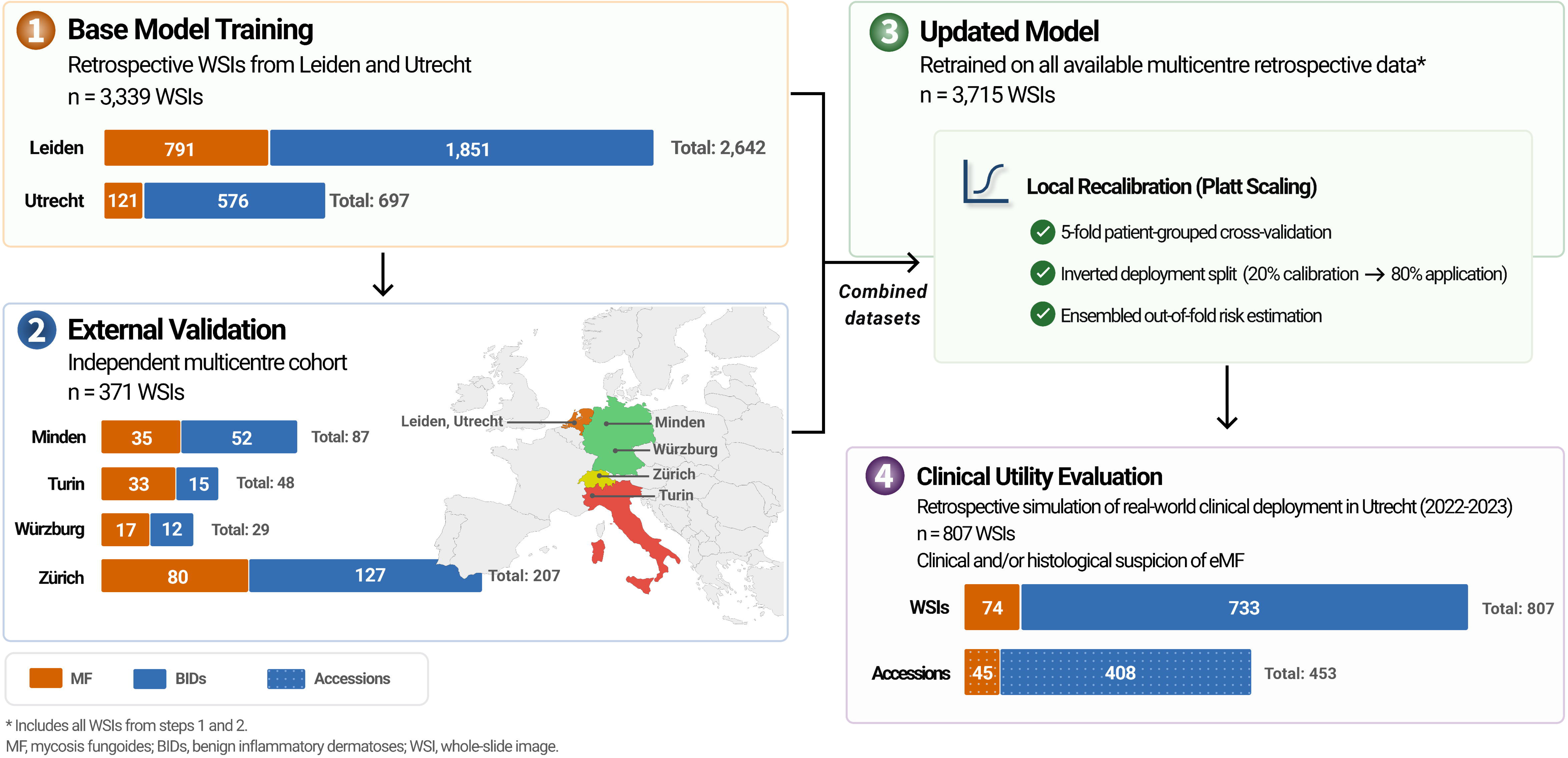
External validation of the base model across four European centres. (A) Receiver operating characteristic (ROC) curves showing stratified performance across four independent external cohorts: Turin (n=48 WSIs), Zurich (n=196), Würzburg (n=29), and Minden (n=87). The thick black line represents pooled external validation performance (AUROC 0·84, 95% CI 0·80–0·88). Individual centre-specific AUROCs (95% CIs) are listed in the legend. The diagonal dashed line indicates random discrimination (AUROC 0·50). (B) Predicted probability distributions in the pooled external cohort (n=371 WSIs). Violin plots show the distribution of predicted probabilities for MF (n=167) and BIDs (n=204), with internal dashed lines indicating the median and interquartile range (IQR). Median predicted probability was substantially higher for MF than for BID (0·97 [IQR 0·84–1·00] vs 0·60 [0·30–0·81]; Mann– Whitney U test p<0·0001). Individual predictions are overlaid as jittered points. Whereas MF predictions clustered tightly near the upper end of the probability spectrum, BID predictions showed broader dispersion. This asymmetric uncertainty pattern indicates strong probabilistic separation, with diagnostic ambiguity concentrated predominantly within the benign inflammatory differential. (C) LOESS-smoothed calibration plot for pooled WSI-level external validation. The solid black line represents the locally estimated scatterplot smoothing (LOESS) calibration curve, with the shaded grey area showing the 95% confidence interval derived from 1000 bootstrap iterations. The diagonal dashed line indicates perfect calibration. The plot demonstrates the agreement between model-predicted probabilities and observed proportions of MF in the pooled external cohort.

Predicted probabilities differed substantially between diagnostic groups in the pooled external cohort (n=371 WSIs; Mann–Whitney U test p<0·0001). Median predicted probability was higher for MF than for BIDs (0·97 [IQR 0·84–1·00] vs 0·60 [0·30–0·81]) (Figure 2B). MF predictions clustered tightly near the upper end of the probability spectrum, whereas BID predictions showed broader dispersion, reflecting greater heterogeneity among inflammatory mimics.

Although partial overlap was observed in the intermediate-to-high probability range, the distributions remained well separated overall. This asymmetric uncertainty pattern suggests that diagnostic ambiguity was concentrated predominantly within the benign inflammatory differential rather than among malignant cases. Pooled WSI-level calibration analysis showed moderate miscalibration, with systematic overprediction and overconfident predictions (calibration slope 0·68; intercept −1·46), despite preserved discrimination (Figure 2C).

### Reader study

In a blinded reader study (n=171 WSIs; 78 MF, 93 BIDs), the MIMIC model was compared with 11 (dermato-)pathologists using HCE slides only, without clinical information. The readers provided ordinal diagnostic confidence scores, which were converted to probabilistic estimates for performance evaluation. The MIMIC model demonstrated superior discriminative performance compared with the 11 (dermato-)pathologists. The model achieved an AUROC of 0·87 (95% CI 0·81–0·91), exceeding both the mean pathologist performance (AUROC 0·79 ± 0·03) and the best-performing individual reader (AUROC 0·83; Figure 3A). Paired performance analysis showed that MIMIC operated at a higher sensitivity but lower specificity than the average (dermato-)pathologist, while maintaining comparable balanced accuracy (Figure 3B). This pattern reflects a diagnostic profile favouring sensitivity, consistent with a potential role as a triage or rule-out support tool. Agreement analysis using linearly weighted Cohen’s κ demonstrated fair to moderate concordance between MIMIC and individual pathologists (Figure 3C), consistent with known inter-observer variability in the histopathological assessment of eMF.

**Figure 3.**
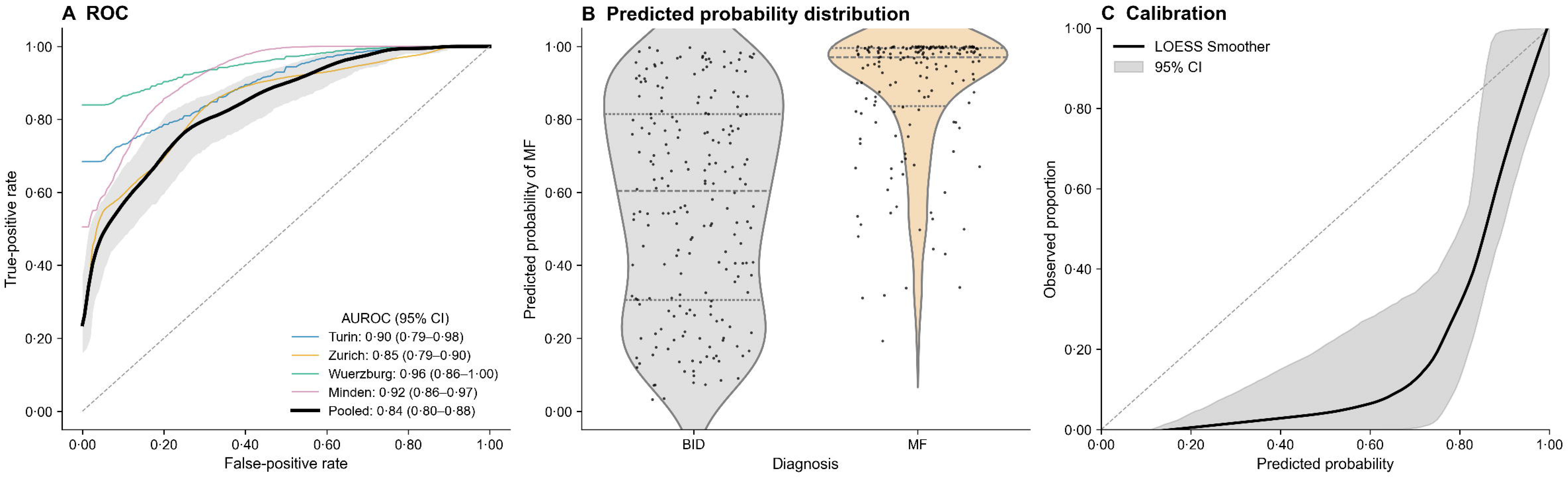
Reader study: diagnostic performance of MIMIC base model compared with 11 (dermato-)pathologists. (A) Receiver operating characteristic (ROC) curve of the MIMIC base model with 95% confidence intervals (shaded area) and operating points of 11 individual pathologists on the same dataset (n=171 slides; 78 MF, 93 BIDs). The model achieved superior discriminative performance (AUROC 0·87, 95% CI 0·81–0·91) compared with the average pathologist (mean AUROC 0·79 ± 0·03), with most readers operating below the model ROC curve. (B) Paired comparison of diagnostic performance between MIMIC and individual pathologists. Points represent individual pathologists and boxplots summarise inter-observer variability. The model demonstrated higher sensitivity but lower specificity compared with the average pathologist, while balanced accuracy was comparable. Error bars for MIMIC represent 95% confidence intervals. (C) Agreement between MIMIC and individual pathologists, measured using linearly weighted Cohen’s κ on a five-point ordinal diagnostic certainty scale. Agreement ranged from fair to moderate across readers, reflecting substantial variability in diagnostic interpretation and alignment between model and human assessments. (D) Performance stratified by case difficulty, defined using a graded response scoring framework based on pathologist agreement. While both MIMIC and pathologists showed reduced performance in diagnostically difficult cases, the decline was substantially greater for pathologists, indicating greater robustness of the model in challenging diagnostic scenarios.

**Figure 4.**
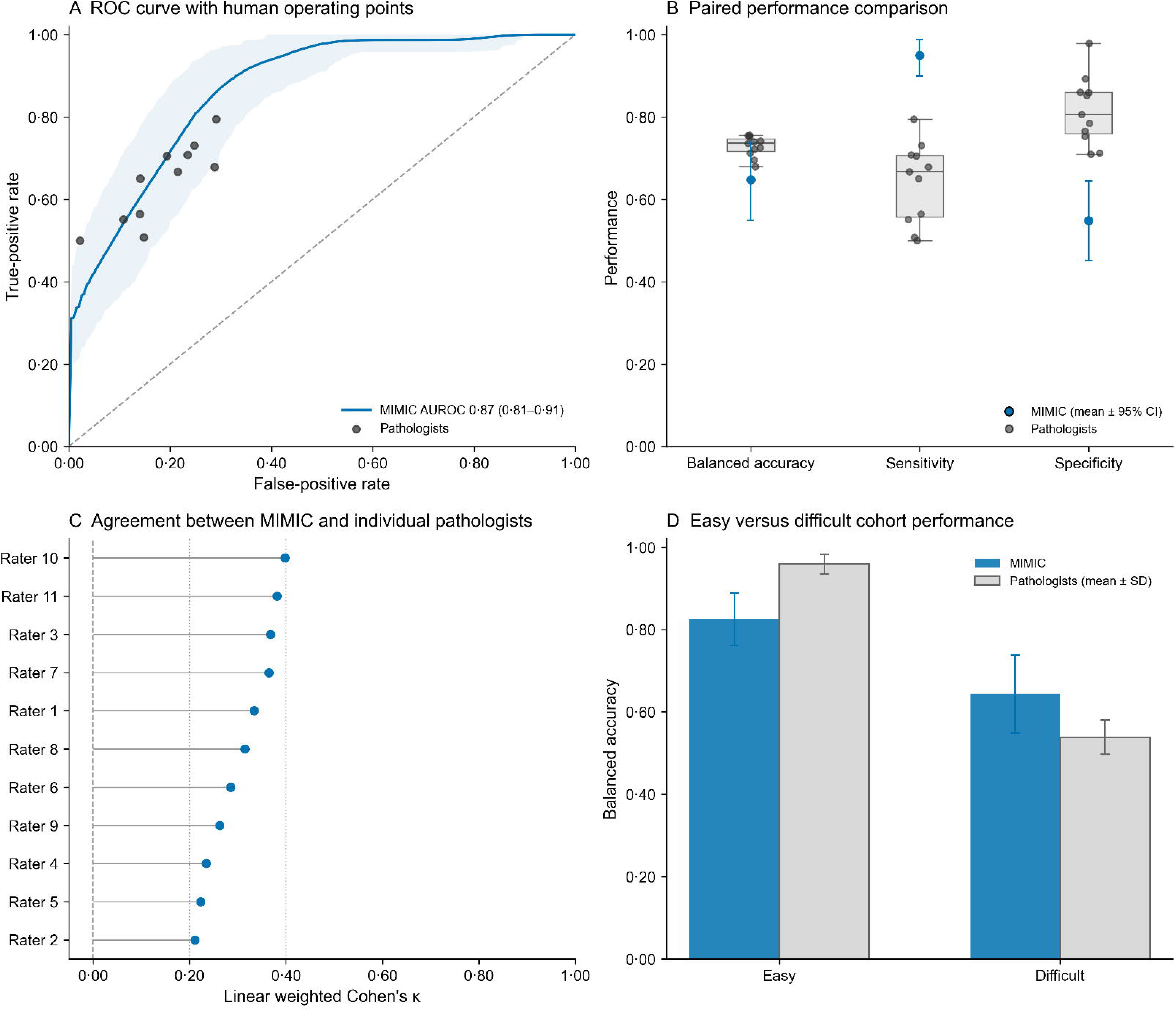
Calibration, discrimination, and risk distribution ( updated model) in the consecutive Utrecht clinical utility cohort. Performance of the updated model in the consecutive Utrecht clinical utility cohort (2022-2023), evaluated at the accession level following max-pooling of WSI predictions. The primary analysis is restricted to MF versus BIDs (n=453 cases; 45 MF cases). (A) Smoothed calibration plot using locally estimated scatterplot smoothing (LOESS) with 95% confidence intervals derived from bootstrapping (1000 iterations). The original model demonstrated substantial miscalibration, with systematic overestimation of risk reflected by a negative calibration intercept (α -2·53) and a slope below unity (β 0·53), indicating overconfident predictions. Prevalence-adjusted intercept scaling using an MF-specific historical accession-level prevalence anchor partially corrected calibration-in-the-large but did not resolve slope miscalibration. The Platt scaled model substantially improved overall calibration, correcting both the intercept (α 0·18) and slope (β 1·18) to more closely align with the ideal diagonal. Rug plots show the distribution of predicted probabilities for MF (top) and BID cases (bottom). (B) Receiver operating characteristic curve with bootstrapped 95% confidence intervals. The model demonstrated good discriminative performance with an AUROC of 0·87 (95% CI 0·81– 0·92), indicating accurate ranking of cases between MF and BIDs despite initial miscalibration of absolute risk estimates. (C) Distribution of predicted probabilities stratified by outcome using the Platt scaled predictions. Violin plots with overlaid data points illustrate the separation between MF and BID cases at the accession level. The dashed blue horizontal line indicates the designated triage threshold of 0·04. Overall, with Platt scaling applied, the model achieves better calibrated absolute risk estimates while retaining its clinically useful ranking performance, effectively supporting its use as a triage tool using the established threshold.

To assess performance under varying levels of diagnostic difficulty, cases were stratified into easy and difficult cohorts based on a graded response scoring framework derived from (dermato-)pathologist agreement. Both MIMIC and humans showed reduced performance in the difficult cohort; however, the decline was substantially greater among humans. While (dermato-)pathologists exhibited a marked drop in balanced accuracy in difficult cases, MIMIC showed a more moderate reduction, indicating greater robustness of the model in diagnostically challenging scenarios (Figure 3D).

### Updated model: temporal deployment validation (accession level-evaluation)

The updated model, retrained on the combined multicentre dataset, was evaluated in the consecutive Utrecht clinical utility cohort. While predictions were generated at the WSI level, evaluation was performed at the accession level by aggregating WSI-level predictions using max-pooling. The primary analysis was restricted to MF versus BIDs, reflecting the intended use of the model as a triage tool for eMF. In this setting (n=453 accessions; 45 MF), the updated model demonstrated good discriminative performance with an AUROC of 0·87 (95% CI 0·81– 0·92). A secondary analysis including all CTCL subtypes (n=486 accessions; 78 CTCL) demonstrated reduced performance (AUROC 0·81; 95% CI 0·76–0·85). This decrease was expected, as non-MF CTCL subtypes were not represented in the training data.

#### Calibration and recalibration

In the primary analysis restricted to MF versus BIDs, evaluation of the original model at the case level revealed substantial miscalibration in the clinical utility cohort. The model systematically overestimated risk, yielding a calibration intercept (α) of −2.53 (95% CI −2·98 to −2·09) and a calibration slope (β) of 0·53 (0·39 to 0·66), indicating overconfident predictions. This miscalibration primarily reflects spectrum bias (a form of selection bias): because the clinical utility cohort is explicitly restricted to a priori suspect cases, the BID cohort consists of “hard mimickers.” This creates a fundamentally different, higher-risk case-mix distribution compared to the broader population seen during model training.

Initial attempts at prevalence-based recalibration, shifting the predictions to account for baseline prevalence differences, proved insufficient. While this adjustment improved calibration-in-the-large by shifting the intercept to −2·00 (−2·39 to −1·61), the calibration slope remained entirely unchanged at 0·53 (0·39 to 0·66). These findings confirmed that the temporal miscalibration was not solely driven by a prevalence shift, but rather by the altered overall probability distribution associated with these highly ambiguous, suspect cases.

To effectively address this spectrum bias, we applied Platt scaling utilizing stratified 5-fold cross-validation on the consecutive utility cohort. This approach successfully recalibrated the model by adjusting the predicted probabilities across the entire distribution, bringing both the intercept (α 0·18, 95% CI −0·41 to 0·77) and the slope (β 1·18, 95% CI 0·88 to 1·48) into much closer alignment with the ideal diagonal. Overall, while the model’s strong discriminative performance and risk ranking capabilities were inherently preserved, Platt scaling successfully restored the reliability of the absolute risk estimates, validating their use for subsequent decision curve analysis and triage threshold determination.

### Clinical utility evaluation (Platt scaled, updated model)

Unlike base-model development and geographic validation, which were performed at the WSI level, the updated model was evaluated in the Utrecht 2022-2023 cohort at the accession level by max-pooling WSI-level predictions within each pathology accession.

Clinical utility was evaluated using the Platt scaled (recalibrated) updated model on a strictly held-out clinical utility cohort of consecutive cases. In the primary analysis restricted to MF versus BIDs (n=453 cases; 45 MF cases), decision curve analysis demonstrated that the model-assisted strategy provided a consistently higher net benefit compared with both the “test all” (current standard-of-care) and “test none” strategies across the clinically relevant threshold range of 0·01–0·10 (Figure 5).

**Figure 5.**
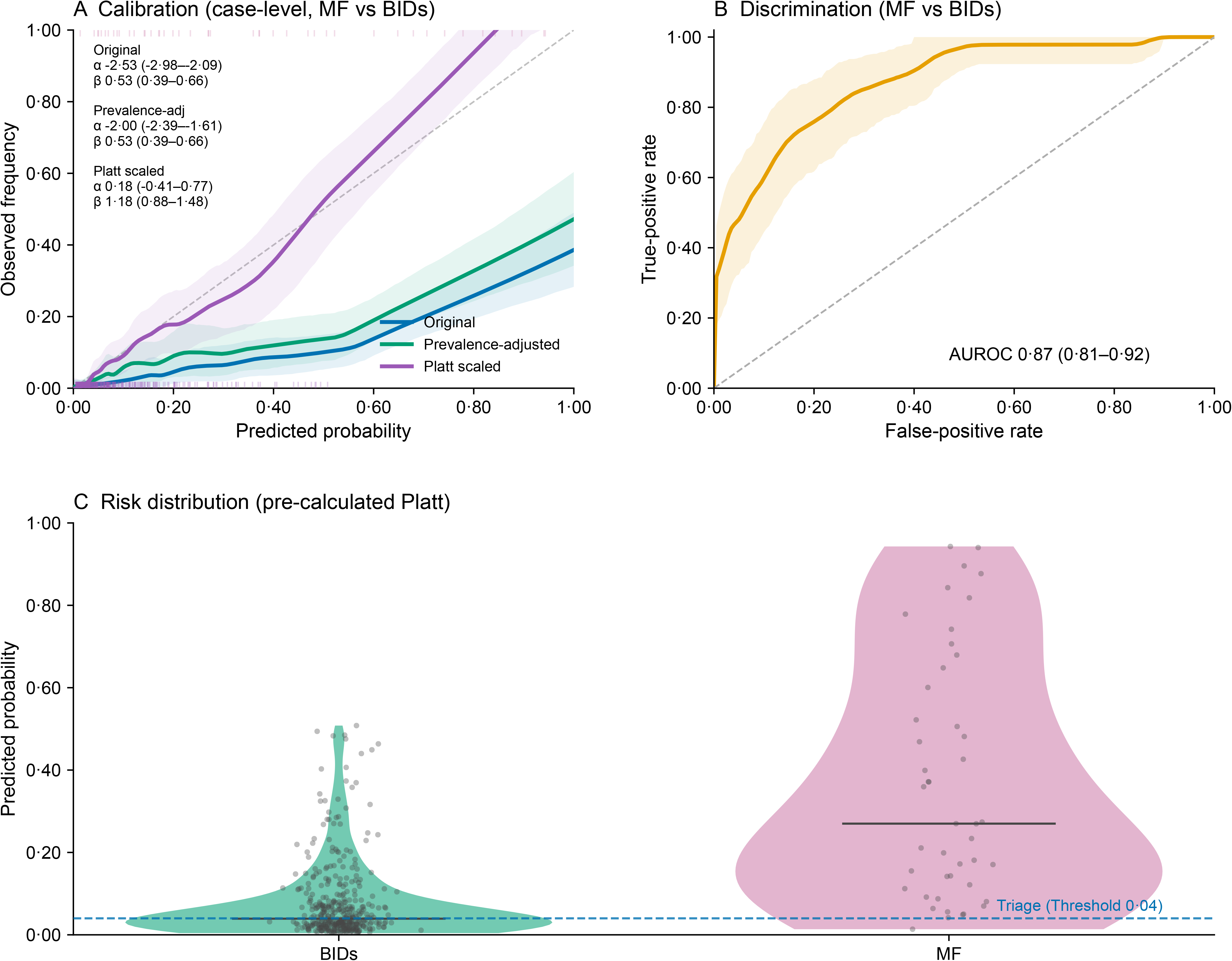
Decision curve analysis and net reduction in unnecessary interventions (prevalence-adjusted, updated model) Clinical utility of the prevalence-adjusted updated model evaluated at the accession level following max-pooling of WSI predictions. The primary analysis is restricted to mycosis fungoides (MF) versus BIDs (n=453 cases; 45 MF cases). (A) Decision curve analysis showing net benefit across threshold probabilities from 0·01 to 0·60. The model-assisted strategy demonstrates positive net benefit relative to both “test all” and “test none” strategies across the clinically relevant threshold range. The shaded region indicates thresholds considered clinically plausible for triage decision-making (0·01–0·10). (B) Net reduction in unnecessary interventions per 100 cases relative to a “test all” strategy across the same threshold range. The Platt scaled model reduces unnecessary interventions while maintaining high sensitivity. The red step-function (right y-axis) indicates the cumulative number of missed MF cases (false negatives) as the decision threshold increases. At a threshold of 0·04, the model achieves a reduction of 40 unnecessary interventions per 100 cases while missing 1 of 45 MF cases. Increasing the threshold improves efficiency but results in a higher number of missed MF cases, illustrating the trade-off between diagnostic safety and resource reduction. The circular markers indicate the selected safety-oriented operating point for triage deployment. (C) Integrated clinical workflow for MF diagnosis incorporating AI-assisted triage. Proposed clinical workflow integrating the MIMIC system into the routine diagnostic pathway for suspected MF. Following an initial skin biopsy with suspicion of MF, cases are evaluated by the MIMIC AI triage system, which stratifies them into three distinct confidence categories. Cases classified as “MF - high confidence” (2·9% of cases) achieve a positive predictive value (PPV) of 100%; for these, further ancillary testing is considered optional before proceeding to multidisciplinary team (MDT) review for clinicopathological correlation and definitive MF diagnosis. The majority of cases fall into the “Grey zone” (51·7%), where ancillary testing is considered essential prior to MDT review to accurately differentiate between MF and no MF. Cases classified as “No MF - high confidence” (45·5% of cases) achieve a negative predictive value (NPV) of 99·5%; for this group, ancillary testing is deemed not necessary, allowing them to safely bypass MDT review and be diagnosed as “no MF”. This workflow illustrates the intended role of MIMIC in streamlining the diagnostic process: prioritizing specialist resources and ancillary testing for ambiguous or high-risk cases, while safely reducing unnecessary downstream investigations and specialist workload for patients with a very low suspicion of MF.

To evaluate the clinical utility of the model as an initial triage tool, a threshold sensitivity analysis was performed. Given the clinical imperative to avoid false-negative diagnoses in eMF, threshold selection prioritised diagnostic safety. While a negative predictive value (NPV) of 95% might be considered acceptable in some contexts, a highly conservative target of >99% NPV is desirable to safely reduce unnecessary benign work-up without compromising patient safety. At a strict decision threshold of 0·04, MIMIC achieved an NPV of 99·5% and a sensitivity of 97·8%, missing only a single case of MF. Simultaneously, the model yielded a specificity of 50·2%, effectively identifying half of all BIDs as low-risk. Implementation at this threshold corresponded to a net reduction of 39·9 unnecessary interventions per 100 cases relative to a “test all” strategy, suggesting substantial reduction in ancillary testing and expert multidisciplinary review while preserving near-complete diagnostic sensitivity. Increasing the threshold to 0·10 improved specificity to 77·7% and increased net reduction to 50·15 interventions per 100 cases, but severely reduced sensitivity to 77·8%, resulting in 10 missed MF cases. Accordingly, a threshold of 0·04 was selected as the optimal safety-oriented operating point for prospective triage deployment.

In terms of algorithmic fairness, sensitivity analyses at the 0·04 threshold demonstrated generally equitable performance across patient sex, age, anatomical site, and slide age, with balanced accuracy estimates ranging from approximately 0·62 to 0·79 across strata (Supplementary Figure 3). Within anatomical-site subgroups, discrimination was lower in the upper extremity subgroup (N = 155, AUROC ∼0·72) relative to the overall cohort. Performance estimates for cases with undocumented anatomical site were imprecise because of the small sample size (N = 14), resulting in wide confidence intervals. Documentation of Fitzpatrick skin type was extremely limited (<5%), precluding meaningful analysis.

In the secondary exploratory analysis including all CTCL subtypes (n=486 cases; 78 CTCL cases), clinical utility was substantially attenuated. Applying the previously established threshold of 0·04 to predictions calibrated by the new CTCL-specific Platt scaler preserved sensitivity at 100%, but specificity plummeted to 7·6%. This severe drop in specificity at a fixed threshold reflects the upward shift in calibrated probabilities across the benign cohort caused by fitting the Platt scaler to a higher-prevalence, variant-inclusive data distribution. This corresponded to a marginal net reduction of only 6·4 unnecessary interventions per 100 cases relative to a “test all” strategy, demonstrating a near-complete loss of triage efficiency. Attempting to improve efficiency by increasing the threshold resulted in an unacceptable drop in safety. For example, at a threshold of 0·10 on this newly calibrated scale, specificity improved to 51·0%, but sensitivity fell to 91·0%, resulting in 7 missed CTCL cases (the upward calibration shift having artificially pushed several previously missed MF cases above this threshold). Diagnostic review of these false negatives revealed a predominance of subtypes not represented in the primary training data, specifically folliculotropic MF (FMF; n=2), Sézary syndrome (n=2), and lymphomatoid papulosis (LyP; n=1), alongside a minority of classic MF cases (n=2). These findings illustrate the critical disconnect between statistical discrimination and clinical utility: while the model retains the ability to rank generalized CTCL risk, its optimal deployment as a safe and efficient triage tool remains strictly confined to its intended target population of classic eMF.

### Error analysis

To characterize model behaviour across a broader spectrum of low-confidence predictions, a detailed qualitative audit of the clinical utility cohort was performed by two pathologists (TDO and ASC) for the false-negative cases below a 0·10 threshold (n=10), including the single missed MF case at the 0·04 operational threshold.

This analysis revealed three cases with atypical presentation of MF, missing epidermotropism. Seven other cases were morphologically consistent with eMF, while LRP heatmaps and top-12 tiles revealed atypical lymphocytes at the dermo-epidermal junction; model failure for these cases remains unexplained.

Next, for the false-positives above a 0·40 threshold, 10 cases were audited. All cases (10/10) were morphologically regarded as (slightly) suspicious for eMF, where indeed IHC and clinical correlation were essential to derive the BID diagnosis. These included cases classified as atopic dermatitis (some treated with JAK-inhibitor or biologics), lichen striatus, and lichenoid dermatitis with post-inflammatory hyperpigmentation. Notably, for two cases, there was no known clinical follow-up, reducing the value of the diagnostic label, and for one other case, lesions resolved at follow-up, yielding no definitive diagnosis. Importantly, 2 out of 10 cases are provisionally considered to be misidentified cases of eMF, pending further investigations. Extended details for all the audited cases are described in Supplementary Table 2.

## Discussion

In this multicentre study, we developed, externally validated, and evaluated the clinical utility of MIMIC, a pathology foundation-model-based deep learning system for histological triage of eMF within a cohort of skin biopsies already selected for clinical and/or histological suspicion of the disease.

By safely avoiding ancillary testing in 45·5% of all suspected cases (Rule-out / Tier 1) and fast- tracking 2·9% of highly suspicious cases directly to multidisciplinary review (Rule-in / Tier 3), this AI-assisted workflow is anticipated to yield substantial positive impacts both economically and operationally. In our two-year institutional cohort alone, the absolute reduction of 206 unnecessary workups translates to direct laboratory cost savings of approximately €36,000– €72,000 on IHC panels alone. When indicated, additional TCR analysis carries a standardized national inter-institutional tariff of €861 per analysis in the Netherlands (2026 rates). While the actual financial impact may be lower depending on specific institutional agreements and in-house testing capabilities, averting this molecular testing in 206 low-risk cases undeniably yields additional cost reduction for the healthcare system. These estimates completely exclude the additionally averted costs of specialist multidisciplinary review time. Regarding turnaround time, in our current standard-of-care workflow, processing an IHC marker panel requires an average of 1 workday (with susceptibility for further delays), while a TCR clonality study requires a target maximum of 5 workdays. Bypassing this reflexive ancillary testing inherently eliminates at least 6 workdays of laboratory processing time for nearly half of all suspected cases. Although we do not yet have data on the total time to definitive MF diagnosis at the patient-level, eliminating this 6-day laboratory bottleneck removes a major structural barrier to faster diagnosis, thereby optimising specialist resource allocation.

Our results demonstrate that MIMIC possesses excellent transportability across diverse European centres (mean centre-specific AUROC 0·91) and significantly outperforms the average individual (dermato-)pathologist in a blinded reader study. Crucially, in a real-world, consecutive cohort (Utrecht 2022–2023) with suspicion of eMF, MIMIC achieved a sensitivity of 97·8%, missing only a single case of MF, while using a diagnostic safety-focused threshold of 0·04. Simultaneously, the model yielded a specificity of 50·2%, effectively identifying half of all BIDs as low-risk. These findings suggest that implementing AI at the point of initial HCE review can serve as an adjunct triage tool, potentially allowing a substantial proportion of unnecessary IHC and molecular workups and expert referrals to be safely avoided without compromising diagnostic safety.

The clinical urgency for an objective triage tool is underscored by the profound diagnostic delays characteristic of eMF. Recent international survey data from Beatty et al. revealed that the vast majority of experts and general practitioners acknowledge that the diagnosis of eMF is structurally delayed, primarily due to clinical and histological misclassification as BIDs^16^. Histopathological evaluation remains inherently subjective and is fundamentally reliant on clinicopathological correlation. Indeed, Hadi et al. demonstrated that the availability of clinical photographs significantly improves both the diagnostic accuracy and confidence of dermatopathologists, particularly when ruling out benign inflammatory mimics^17^.

By achieving a global AUROC of 0·87 and exceeding the overall discriminative area of both the mean pathologist (0·79) and the top-performing individual reader (0·83), MIMIC demonstrates that a purely histological, objective deep learning signal can mitigate interobserver variability. Although certain pathologists achieved superior localized efficiency at discrete, implicit operating points on the ROC space, the model provided superior overall discrimination across the continuous threshold spectrum. This continuous performance profile is a critical attribute for a dynamic triage tool, as human readers cannot easily modulate their internal clinical thresholds to maintain high safety margins, particularly in ultra-high sensitivity zones. While this unimodal HCE approach ensures maximum accessibility, scalability, and seamless integration into routine workflows without the hurdle of acquiring perfectly structured metadata, the future integration of clinical photography and patient characteristics into a multimodal foundation model represents the logical next step. By computationally mirroring the multidisciplinary diagnostic process, such multimodal integration could further enhance diagnostic specificity and ultimately provide a holistic, end-to-end diagnostic support system.

Our findings should be contextualised within the rapidly evolving landscape of artificial intelligence applied to MF diagnostics. Recent work by Zhao et al. demonstrated the feasibility of differentiating MF from BIDs using a self-supervised multimodal framework that integrates HCE whole-slide images with structured clinical variables, achieving a macro-balanced accuracy of 0·84 ^18^. However, their reliance on structured clinical metadata limits real-world applicability, as such data are frequently missing or incomplete during routine histological review. Furthermore, their multiclass algorithm was restricted to differentiating MF from only three specific inflammatory conditions (atopic dermatitis, psoriasis, and drug reactions). This restricted scope artificially simplifies the diagnostic challenge; in contrast, MIMIC relies exclusively on unimodal HCE images and encompasses a broad, unselected case-mix of BIDs, which accurately reflects the heterogeneous spectrum of inflammatory mimics encountered in routine clinical triage workflows. By employing rigorous temporal evaluation and prevalence-based recalibration, our approach specifically addresses these generalisation hurdles. Similarly, while preliminary reports of other unimodal HCE models, such as recent conference abstracts by Sharaf et al., have reported high accuracies, these systems were evaluated on heavily restricted sample sizes^19,20^, increasing the risk of overfitting relative to our large-scale, multicentre validation.

Importantly, when evaluating MIMIC under realistic deployment conditions where clinically distinct but histologically overlapping non-MF CTCL entities are inevitably present, we observed an anticipated attenuation in performance (AUROC 0·81). Diagnostic review of these false negatives revealed a predominance of subtypes entirely unrepresented in the primary training data, including folliculotropic MF (*n* = 2), Sézary syndrome (*n* = 2), lymphomatoid papulosis (*n* = 1). This reduction in performance reflects a known mismatch between the model’s predefined training target (classic MF) and the broader morphological spectrum of cutaneous lymphomas, rather than an algorithmic failure within its intended scope. Consequently, these findings underscore that optimal deployment of MIMIC must be strictly confined to its intended target population of classic early-stage patch- and plaque-stage MF, emphasizing the need for a precise definition of intended use when deploying AI systems in heterogeneous disease groups.

The diagnostic pathway for eMF is currently hampered by a median delay of 36 months, largely due to the profound morphological overlap between early malignant infiltrates and BIDs. In tertiary referral centres, this leads to a “defensive diagnostic reflex” where IHC and molecular clonality studies are frequently requested for cases with only marginal morphological suspicion. Following robust recalibration via Platt scaling to correct for local spectrum bias, we demonstrated through Decision Curve Analysis (DCA) a consistent positive Net Benefit across a clinically relevant range of risk thresholds (1–10%). This indicates that MIMIC could effectively de-bulk the “equivocal pile” of biopsies, ensuring that specialized resources and ancillary tests are prioritised for the most diagnostically challenging patients.

The modest impact of initial attempts at prevalence-based adjustment (anchored to a historical prevalence of 0·130) suggests that temporal miscalibration was not explained by prior probability shift alone; the persistence of a severe slope miscalibration (β = 0·53) indicated broader distributional differences (spectrum bias) between the retrospective training data and the highly suspect real-world deployment cohort. We therefore applied Platt scaling to the Utrecht cohort to restore reliable absolute risk estimates for our decision curve analysis. However, because Platt scaling mathematically fits to a local probability distribution rather than a single prevalence metric, this calibration profile may not perfectly transport to institutions with different referral patterns. Consequently, adopting hospitals will likely need to evaluate local calibration or compute their own Platt scaling parameters before relying on absolute risk thresholds for triage. Similarly, while the 0·04 safety threshold successfully reduced unnecessary workups locally, future prospective multicentre studies must verify if this threshold requires adjustment to reflect varying institutional case-mixes. Accordingly, MIMIC should currently be interpreted as a high-sensitivity ranking and triage tool, not as a universally calibrated probability estimator, and its outputs are intended to be used strictly in conjunction with clinical and histopathological judgement.

### Limitations

Despite its strong performance, this study has several limitations. First, while our consecutive clinical utility set mirrors a real-world workflow, its restriction to highly suspicious cases introduced severe spectrum bias that necessitated local Platt scaling. Consequently, the absolute risk estimates and our optimal 0·04 triage threshold may not safely transport to other institutions without local recalibration. Second, MIMIC currently operates as a single-modality system, lacking integration with clinical data (e.g., lesion distribution or duration) and TCR clonality results. Crucially, while the model identifies nearly all classic eMF cases, our data shows it loses safe triage utility when applied to non-MF CTCL variants (e.g., Sézary syndrome, FMF), confirming its use must be strictly confined to classic eMF. Third, documentation of Fitzpatrick skin type was extremely limited in both the development and clinical utility cohorts, and patients with darker skin types (IV–VI) were under–represented. External validation in populations with a broader distribution of skin phototypes and outside European centres is therefore required before generalising these findings.

A recognized limitation of our validation design is the reliance on standard-of-care clinicopathological correlation for the BID control group. Because eMF is characterized by an indolent course that closely mimics benign dermatoses, and longitudinal follow-up was not universally available for all control patients, we cannot entirely exclude the possibility that a small fraction of the BID cohort harbours undiagnosed, early-evolving MF. Indeed, our error analysis exposed two cases provisionally considered to be misidentified cases of eMF, pending full investigations. This potential label noise would be expected to bias our performance estimates towards the null, such that the true discriminative ability of the model for unequivocally benign inflammatory dermatoses is likely higher than reported. However, because advanced ancillary testing was utilized for close mimics in our cohorts, and this pattern of imperfect labelling closely mirrors the real-world diagnostic constraints under which an AI triage tool must operate, we believe that the reported performance metrics provide a conservative yet clinically realistic estimate of the model’s utility.

While we applied Platt scaling to capture the exact clinical case-mix and accurately anchor our Decision Curve Analysis, these real-world distributional dynamics were by definition unavailable for the retrospective, curator-selected external validation cohorts from the CLIDIPA registry. Consequently, our local Platt-scaled recalibration was strictly optimized for the clinical utility evaluation, and regional variations in baseline threshold utility across the external European centres remain to be fully characterized in prospective, unmanipulated screening workflows. Future prospective deployment studies should capture centre-specific case-mix distributions to compute localized Platt scaling parameters and determine whether these further optimize regional triage efficiency.

### Future Perspectives and Conclusion

MIMIC is uniquely positioned to function as a highly scalable and resource-efficient initial triage step within the pathology workflow. While emerging molecular classifiers offer promising sensitivity and specificity for eMF^21^, these ancillary tests are resource-intensive, require additional tissue, and are not universally accessible. By leveraging existing digitised HCE slides, a highly sensitive AI triage model can optimally identify which ambiguous cases actually warrant these expensive molecular or immunohistochemical investigations. This approach optimises laboratory resource allocation at negligible marginal cost, effectively reducing the economic and logistical burden of unnecessary interventions.

From an implementation perspective, MIMIC is intended as a computer–assisted diagnostic support tool integrated into existing digital pathology systems, rather than an autonomous diagnostic device. It is designed as a high-sensitivity risk-stratification aid to be used at the moment of initial clinical or histological suspicion, before committing to costly downstream interventions such as ordering IHC panels or expert centre/panel referral. The model is strictly intended for use by trained pathologists in centres with digital WSI infrastructure; users require basic familiarity with AI–assisted decision support and local calibration procedures. Furthermore, in routine use, WSIs with severe artefacts, fading, or a missing epidermis should be rescanned or excluded before model inference, as these conditions were not used for training. Local deployment will additionally require centre–specific threshold selection, periodic re–audit of performance, and alignment with institutional policies and applicable medical device regulations.

Looking forward, histological decision support systems like MIMIC could be integrated into a comprehensive, end-to-end digital ecosystem for CTCL management. This pathway could begin upstream with predictive machine learning models that flag high-risk patients based on their dermatological history and electronic health records, such as previous treatment failures or multiple inconclusive biopsies^22^. Flagged cases could then be evaluated using multimodal convolutional neural networks for clinical and dermoscopic imaging^23^, seamlessly followed by MIMIC for objective, ’zero-cost’ histological triage. Once an eMF diagnosis is established, prognostic deep learning algorithms applied to the very same routine HCE slides could instantly predict the individual risk of disease progression to advanced stages, enabling early risk-stratification^24^. Finally, for patients with confirmed MF, novel 3D AI models (such as mSWAT-Net) can automate the calculation of skin tumour burden to longitudinally monitor disease progression and treatment efficacy^25,26^. Working synergistically, these tiered AI applications have the potential to fundamentally streamline the entire diagnostic, prognostic, and monitoring pathway for patients with CTCL.

In conclusion, MIMIC represents a significant step towards the objective, risk-stratified triage of eMF. By providing a high-sensitivity filter at the earliest stage of the pathology workflow, the model offers a viable solution to reduce diagnostic burden and expedite the journey from first biopsy to definitive diagnosis for patients with MF. Future research should focus on the prospective deployment of MIMIC to evaluate its impact on diagnostic turnaround times and cost-effectiveness in routine practice.

## Contributors

### Data sharing

The large Leiden and Utrecht training datasets of HCE-stained WSIs related to this article cannot be made publicly available owing to legal restraints. The external validation datasets and subsets of Leiden and Utrecht data are available to European research groups that demonstrate a long-term commitment to AI-based research in cutaneous lymphomas by becoming a member of the CLIDIPA Registry (https://clidipa.org/). Affiliated sites are invited to submit their study proposals to access these data. Projects eligible to request data from the Registry must focus primarily on using the data as a validation cohort for internally developed AI-models.

## Supporting information

TRIPOD+AI checklist

## Data Availability

The large Leiden and Utrecht training datasets of H&E-stained WSIs related to this article cannot be made publicly available owing to legal restraints. The external validation datasets and subsets of Leiden and Utrecht data are available to European research groups that demonstrate a long-term commitment to AI-based research in cutaneous lymphomas by becoming a member of the CLIDIPA Registry (https://clidipa.org/). Affiliated sites are invited to submit their study proposals to access these data. Projects eligible to request data from the Registry must focus primarily on using the data as a validation cohort for internally developed AI-models.

https://github.com/LUMCPathAI/MIMIC

https://clidipa.org/

## Acknowledgments

This work was supported by an unrestricted Fellowship grant of Stichting Hanarth Fonds in The Netherlands. We would like to acknowledge Rein Willemze and all members of the Dutch Cutaneous Lymphoma Group for establishing the registry of the Dutch Cutaneous Lymphoma Group that served as the basis for the Leiden and Utrecht whole-slide image datasets developed for this study. During the preparation of this work, the author(s) used NotebookLM (powered by Google Gemini) to assist with language review, syntax correction, and improving the general readability of the manuscript text. The author(s) reviewed and edited the output as needed and take full responsibility for the content of the published article.

## Supplementary Material (6)

Supplementary Table 1: Histopathological reaction patterns and specific diagnoses of benign inflammatory dermatoses in the development cohort.

Supplementary Table 2: Detailed qualitative error audit of the updated model for the clinical utility cohort.

Supplementary Figure 1: Technical benchmarking of multiple instance learning architectures and pathology foundation models (base model).

Supplementary Figure 2: Flow diagram of patient selection for clinical utility evaluation.

Supplementary Figure 3: Subgroup analysis of MIMIC diagnostic performance across demographic, clinical, and pre-analytical characteristics.

TRIPOD+AI checklist

---

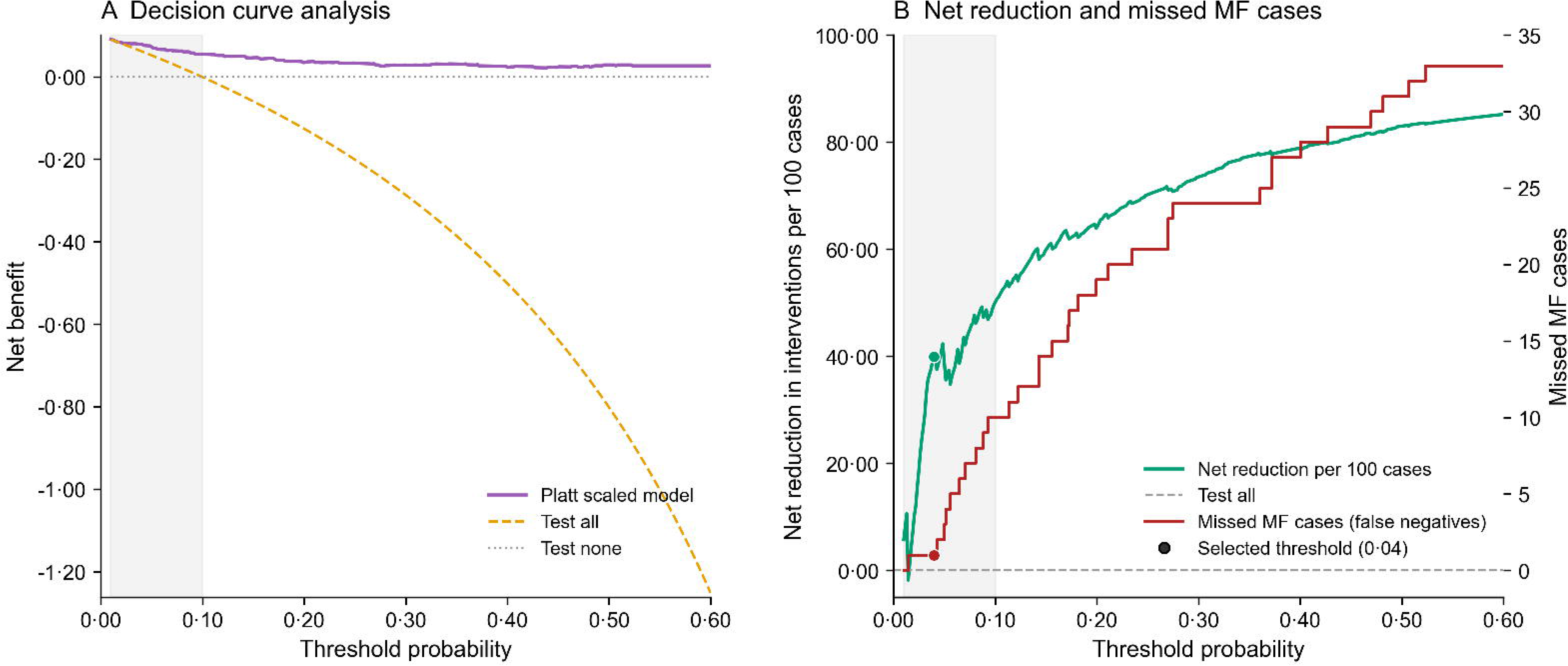

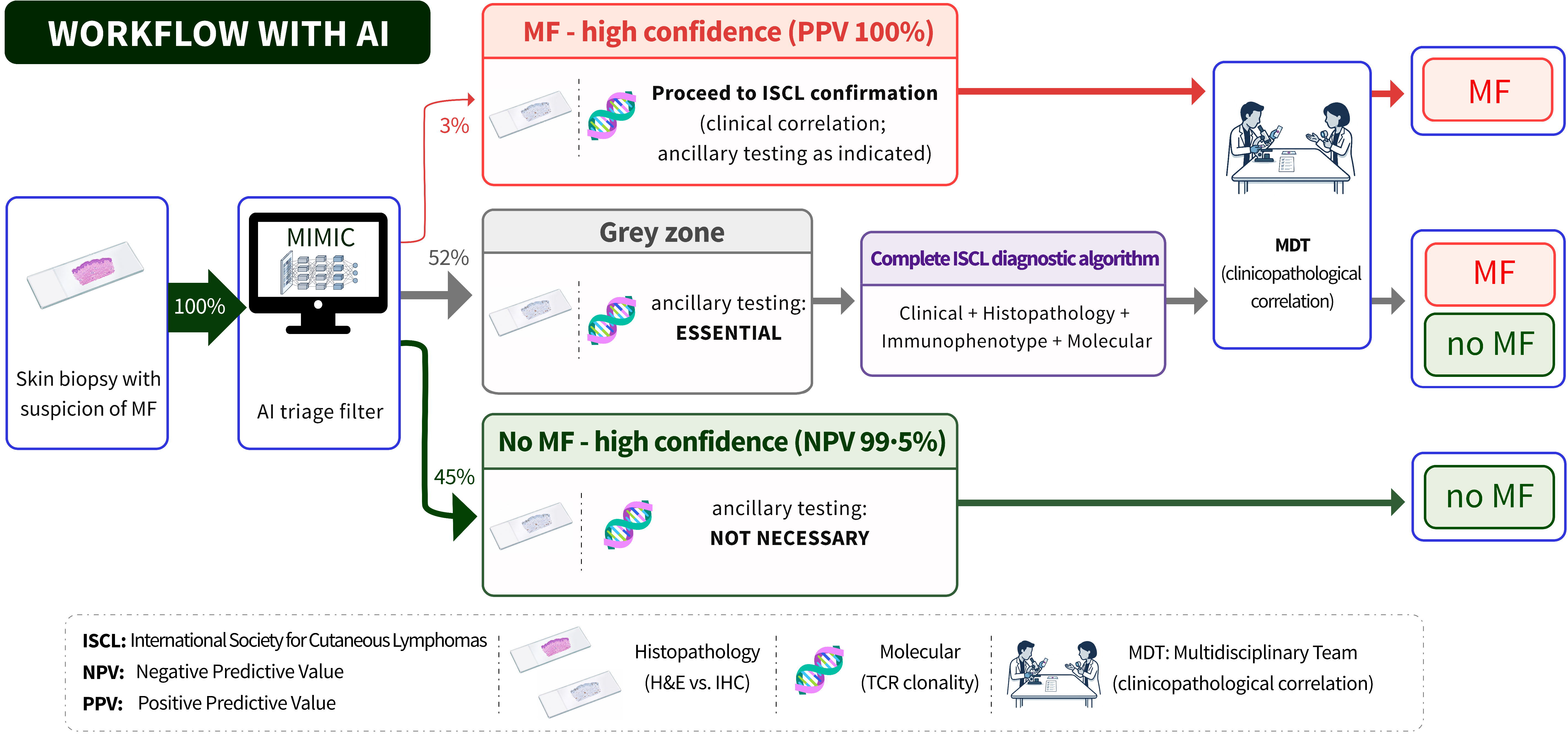

## References

1. Hodak E, Amitay-Laish I. Mycosis fungoides: A great imitator. Clinics in Dermatology 2019; 37(3): 255–67.

2. Scarisbrick JJ, Quaglino P, Prince HM, et al. The PROCLIPI international registry of early-stage mycosis fungoides identifies substantial diagnostic delay in most patients. Br J Dermatol 2019; 181(2): 350–7.

3. Pimpinelli N, Olsen EA, Santucci M, et al. Defining early mycosis fungoides. Journal of the American Academy of Dermatology 2005; 53(6): 1053–63.

4. Hodak E, Geskin L, Guenova E, et al. Real-Life Barriers to Diagnosis of Early Mycosis Fungoides: An International Expert Panel Discussion. American Journal of Clinical Dermatology 2022: 1–10.

5. Doeleman T, Brussee S, Hondelink LM, et al. Deep Learning-Based Classification of Early-Stage Mycosis Fungoides and Benign Inflammatory Dermatoses on HCE-Stained Whole-Slide Images: A Retrospective, Proof-of-Concept Study. J Invest Dermatol 2025; 145(5): 1127–34 e8.

6. Collins GS, Chester-Jones M, Gerry S, et al. Clinical prediction models using machine learning in oncology: challenges and recommendations. BMJ Oncology 2025; 4(1): e000914.

7. Collins GS, Moons KGM, Dhiman P, et al. TRIPOD+AI statement: updated guidance for reporting clinical prediction models that use regression or machine learning methods. BMJ 2024; 385: e078378.

8. Jawed SI, Myskowski PL, Horwitz S, Moskowitz A, Querfeld C. Primary cutaneous T-cell lymphoma (mycosis fungoides and Sézary syndrome): Part I. Diagnosis: Clinical and histopathologic features and new molecular and biologic markers. Journal of the American Academy of Dermatology 2014; 70(2): 205.e1-.e16.

9. Cerroni L. Skin lymphoma: the illustrated guide: John Wiley & Sons; 2020.

10. Moons KGM, Damen JAA, Kaul T, et al. PROBAST+AI: an updated quality, risk of bias, and applicability assessment tool for prediction models using regression or artificial intelligence methods. BMJ 2025; 388: e082505.

11. Scalbert M, Saillard C, Peeters T, et al. Abstract LB174: H-optimus-1: A foundation model for computational histopathology. Cancer Research 2026; 86(8_Supplement): LB174–LB.

12. Brussee S, Valkema PA, Weijer JA, Doeleman T, Schrader AM, Kers J. PathBench-MIL: A Comprehensive AutoML and Benchmarking Framework for Multiple Instance Learning in Histopathology. arXiv preprint arXiv:251217517 2025.

13. Ilse M, Tomczak J, Welling M. Attention-based deep multiple instance learning. International conference on machine learning; 2018: PMLR; 2018. p. 2127–36.

14. Shao D, Chen RJ, Song AH, et al. Do Multiple Instance Learning Models Transfer? In: Aarti S, Maryam F, Daniel H, et al., editors. Proceedings of the 42nd International Conference on Machine Learning. Proceedings of Machine Learning Research: PMLR; 2025. p. 54219--38.

15. Van Calster B, Collins GS, Vickers AJ, et al. Evaluation of performance measures in predictive artificial intelligence models to support medical decisions: overview and guidance. Lancet Digit Health 2025; 7(12): 100916.

16. Beatty P, Molloy K, Porkert S, et al. The diagnosis of early-stage mycosis fungoides: Views held by experts and non-experts in cutaneous lymphoma. JDDG: Journal der Deutschen Dermatologischen Gesellschaft 2025; 23(9): 1094–102.

17. Hadi R, Miller TI, May C, et al. Impact of clinical photographs on the accuracy and confidence in the histopathological diagnosis of mycosis fungoides. Journal of Cutaneous Pathology 2021; 48(7): 842–6.

18. Zhao J, Bai J, Li G, et al. Self-supervised AI system for differentiating mycosis fungoides and benign inflammatory dermatoses. British Journal of Dermatology 2026: ljag129.

19. Sharaf M, Jour G, Smoller B, Giubellino A, Shvartsbeyn M, Park C. 397 Distinction of Mycosis Fungoides From Its Morphologic Mimics Using AI-Powered Analysis of Whole Slide Images. Laboratory Investigation 2026; 106(3, Supplement): 104680.

20. Sharaf M, Shvartsbeyn M, Giubellino A, Park C. Diagnosing mycosis fungoides using AI powered histologic analysis. Blood 2025; 146(Supplement 1): 2570-.

21. Meinel M, Langreder N, Schmitt A, et al. LCK and HOMER1 gene expression-based classifier distinguishes early mycosis fungoides from eczema and psoriasis. medRxiv 2025: 2025.10.18.25338021.

22. Luyten S, Gordon ER, Suhl S, et al. 62310 A Machine Learning Model for Early Detection of Mycosis Fungoides: Distinguishing CTCL from Benign Dermatoses. Journal of the American Academy of Dermatology 2025; 93(3): AB8.

23. Liu Z, Zhang Y, Wang K, Xie F, Liu J. Early diagnosis model of mycosis fungoides and five inflammatory skin diseases based on a multimodal data-based convolutional neural network. British Journal of Dermatology 2025; 193(5): 968–77.

24. Valkema PA, Brussee S, Kersten JM, et al. Deep learning-based prediction of disease progression in early-stage mycosis fungoides based on haematoxylin Camp; eosin-stained whole-slide images. European Journal of Cancer 2025; 229.

25. Wang H, Liu Z, Pan H, et al. A 3-Dimensional-Optimized Artificial Imaging Model for the Skin Tumor Burden Assessment of Mycosis Fungoides. Journal of Investigative Dermatology 2026; 146(1): 55–63.e7.

26. Kittler H. Artificial Intelligence and Skin Disease Scoring: An Opportunity to Reset the Standards. Journal of Investigative Dermatology 2026; 146(1): 9–10.

