## Supplementary Figures for "Histological triage of early-stage mycosis fungoides using a weakly supervised deep learning-based model: a multicentre, external validation, and clinical utility study"

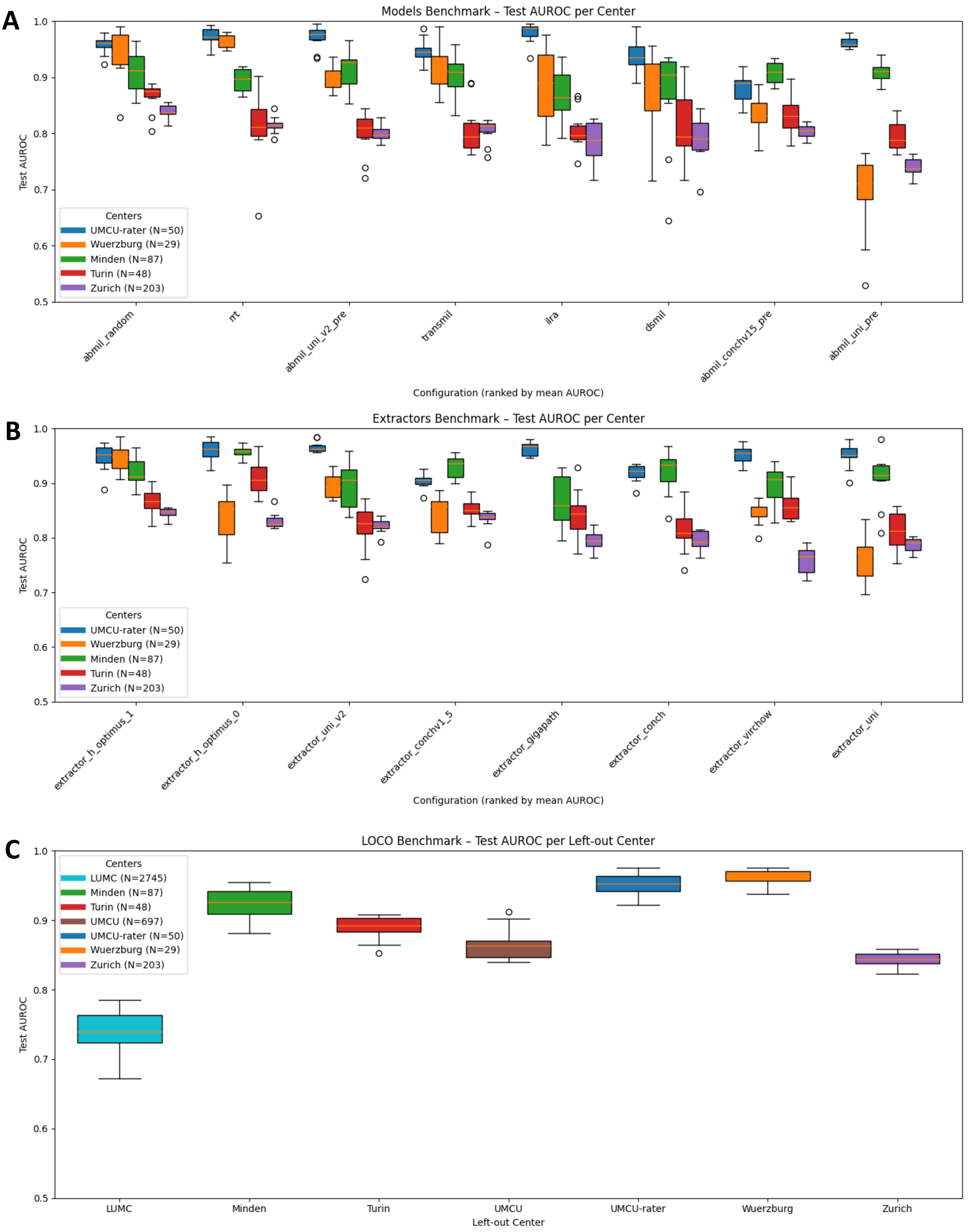


**Supplementary Figure 1: Technical benchmarking of multiple instance learning architectures and pathology foundation models (base model).**

Performance evaluation of various deep learning configurations across four international cohorts. **(A)** Ranking of eight multiple instance learning (MIL) architectures based on mean test AUROC on 10-fold cross-validation. **(B)** Comparison of eight pathology foundation models used as feature extractors, ranked by mean performance. The H-Optimus-1 extractor consistently demonstrated superior robustness. **(C)** Leave-one-centre-out (LOCO) validation results, illustrating the geographic transportability of the MIMIC system. AUROC remained stable across most external sites regardless of the excluded training centre. Exclusion of

Leiden training data evidently reduced AUROC. Boxes represent the interquartile range (IQR), horizontal lines indicate the median, and whiskers extend to $1 \cdot 5 \times$ IQR. Individual points represent outliers. AUROC=area under the receiver operating characteristic curve.

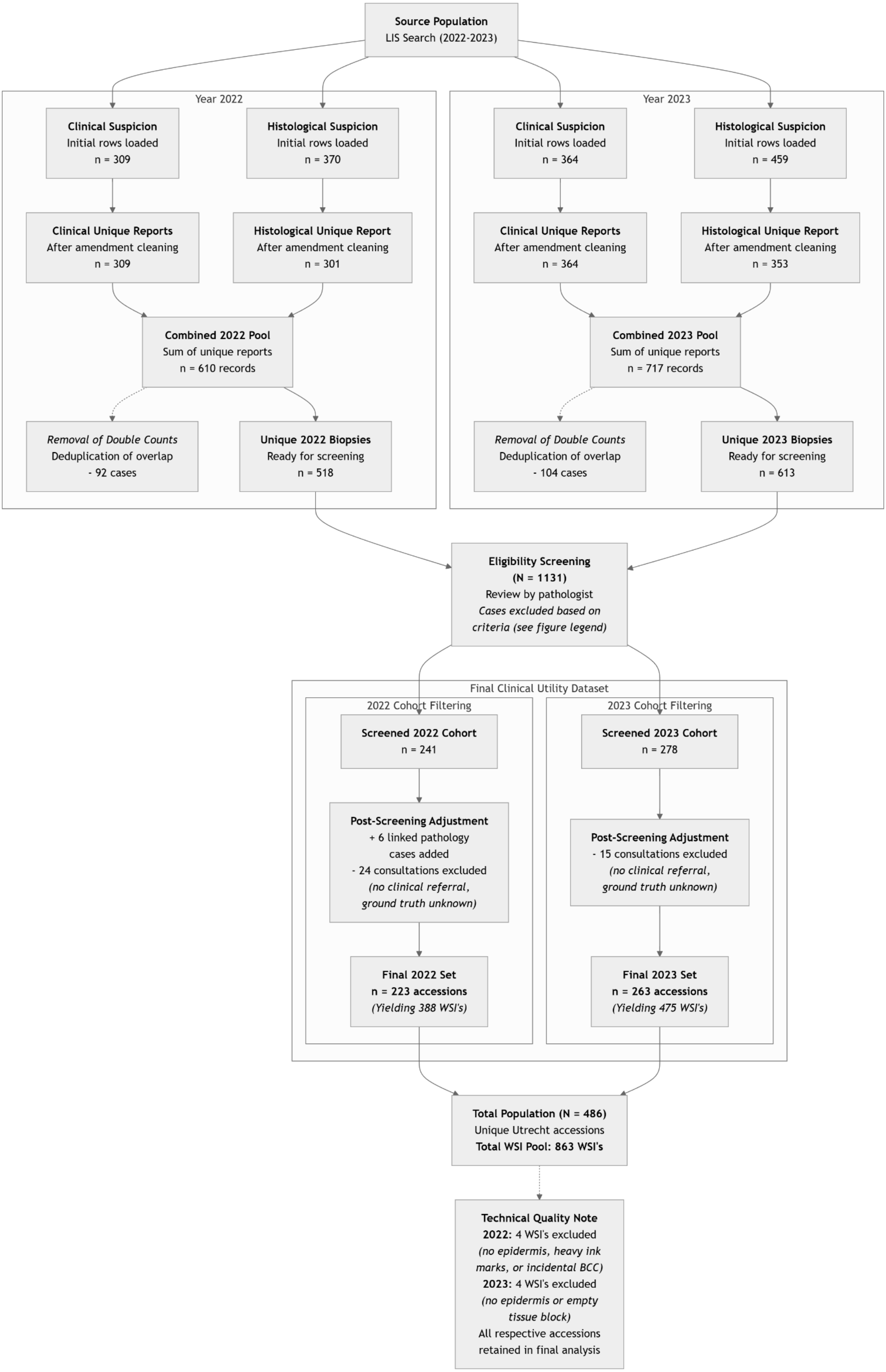
Source Population
LIS Search (2022-2023)
Year 2022
Clinical Suspicion
Initial rows loaded
n = 309
Histological Suspicion
Initial rows loaded
n = 370
Clinical Unique Reports
After amendment cleaning
n = 309
Histological Unique Report
After amendment cleaning
n = 301
Combined 2022 Pool
Sum of unique reports
n = 610 records
Removal of Double Counts
Deduplication of overlap
- 92 cases
Unique 2022 Biopsies
Ready for screening
n = 518
Year 2023
Clinical Suspicion
Initial rows loaded
n = 364
Histological Suspicion
Initial rows loaded
n = 459
Clinical Unique Reports
After amendment cleaning
n = 364
Histological Unique Report
After amendment cleaning
n = 353
Combined 2023 Pool
Sum of unique reports
n = 717 records
Removal of Double Counts
Deduplication of overlap
- 104 cases
Unique 2023 Biopsies
Ready for screening
n = 613
Eligibility Screening
(N = 1131)
Review by pathologist
Cases excluded based on
criteria (see figure legend)
Final Clinical Utility Dataset
2022 Cohort Filtering
Screened 2022 Cohort
n = 241
Post-Screening Adjustment
+ 6 linked pathology
cases added
- 24 consultations excluded
(no clinical referral,
ground truth unknown)
Final 2022 Set
n = 223 accessions
(Yielding 388 WSI's)
2023 Cohort Filtering
Screened 2023 Cohort
n = 278
Post-Screening Adjustment
- 15 consultations excluded
(no clinical referral,
ground truth unknown)
Final 2023 Set
n = 263 accessions
(Yielding 475 WSI's)
Total Population (N = 486)
Unique Utrecht accessions
Total WSI Pool: 863 WSI's
Technical Quality Note
2022: 4 WSI's excluded
(no epidermis, heavy ink
marks, or incidental BCC)
2023: 4 WSI's excluded
(no epidermis or empty
tissue block)
All respective accessions
retained in final analysis

**Supplementary Figure 2: Flow diagram of patient selection for clinical utility evaluation.** BCC: basal cell carcinoma; LIS: Laboratory Information System; MF: Mycosis Fungoides; WSI: Whole Slide Image.

**Eligibility Screening Inclusion Criteria.* Skin biopsy reports from an automated LIS search were included in the Clinical Suspicion screening cohort when these criteria were met: the Clinical Details report section should contain an MF, FMF, CTCL, Sezary syndrome, or ‘lymphoma’ diagnostic query from the submitting clinician. The generic lymphoma query was accepted for patients presenting with cutaneous macules, papules, patches, and/or plaques. Skin biopsy reports from the automated LIS search were included in the Histological Suspicion screening cohort when the Microscopy section contained either CD3 or CD4 stain reporting and the Diagnosis text contained MF, FMF, CTCL, Sezary syndrome or ‘lymphoma’ terminology.

**Eligibility Screening Exclusion Criteria:* Biopsy reports were excluded upon manual review by a (dermato-)pathologist based on pre-specified clinical and histological criteria to isolate early-stage MF suspicion. Specific exclusion reasons included: non-skin material (e.g., lymph nodes, bone marrow, and soft tissue specimens); diagnostic queries indicating alternative malignancies or specific non-MF entities (e.g., intravascular lymphoma, lymphomatoid reaction, B-cell proliferations without full T-cell panel, histiocytosis); presenting (subcutaneous) nodules or tumours with differential diagnoses including melanoma, basal cell carcinoma, plasmacytoma, Merkel cell carcinoma, cutaneous metastasis, skin adnexal tumour, sarcoma, atypical fibroxanthoma, atheroma cyst, granuloma, hidradenitis suppurativa, dermatofibroma, lipoma, pseudolymphoma, tumour-stage MF, or infectious Leishmaniasis; and a history of lymphoma without a concurrent active clinical query or suspicion directed at MF.

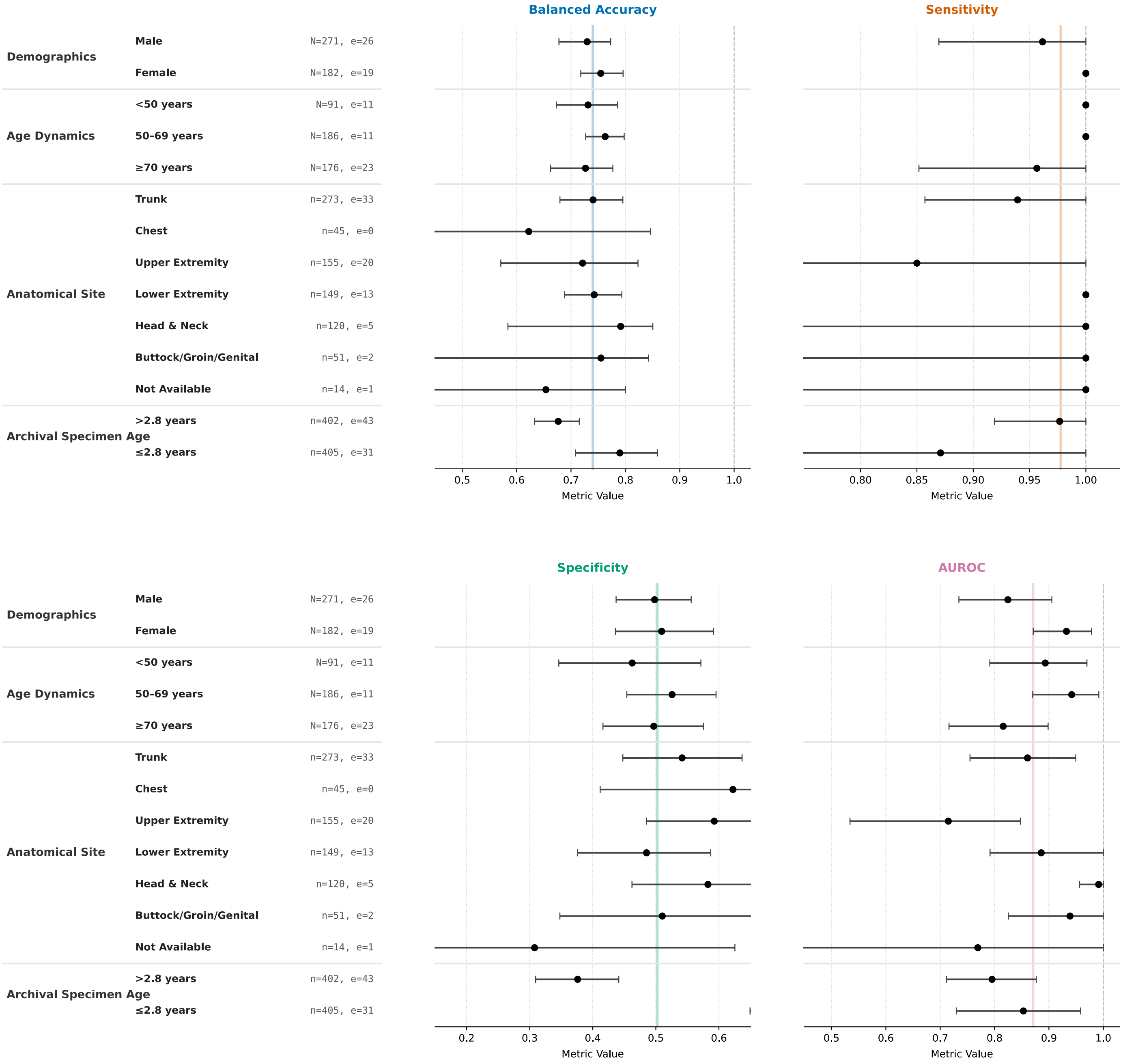

**Supplementary Figure 3. Subgroup analysis of MIMIC diagnostic performance across demographic, clinical, and pre-analytical characteristics.**

Stratified performance metrics of the updated MIMIC model evaluated on the clinical utility cohort ($N = 453$ independent pathology accessions; $n = 807$ WSIs). Performance is reported across four statistical dimensions: Balanced Accuracy, Sensitivity, Specificity, and Area Under the Receiver Operating Characteristic curve (AUROC). To account for data clustering (multiple historical slides nested within a single diagnostic episode), WSI-level predictions were aggregated to the accession level using a max-pooling operator. A case was flagged as high-risk if the predicted probability of any underlying WSI met or exceeded the pre-specified safety threshold of $0 \cdot 04$. Anatomical Site (biopsy location) and Archival Specimen Age (slide age) were analysed at the WSI level ($n = 807$), while patient sex and patient age were evaluated at the aggregated accession level ($N = 453$).

Solid circular markers represent point estimates for each distinct stratum. Horizontal error bars indicate 95% confidence intervals (CIs) calculated via a cluster-robust percentile bootstrap resampling procedure (1,000 iterations), resampling strictly at the patient accession level to account for intra-patient correlation. Vertical coloured lines represent the unstratified baseline performance of the entire validation cohort. Vertical dashed grey lines denote a metric value of $1 \cdot 0$. Tabular notation on the left axis indicates the sample size composition for each stratum, where $N$ (or $n$) represents the total number of units within the subgroup, and $e$ indicates the true event count (confirmed MF cases). Archival specimen age cohorts were dichotomized around the sample median ($2 \cdot 8$ years). Bootstrap confidence intervals for the AUROC metric are omitted for subgroups with insufficient class representation ($e < 5$; e.g., Chest, Buttock/Groin/Genital) where sparse data limited stable estimation.
